# Genetic testing in cancer: a systematic review of patient reported measures

**DOI:** 10.1101/2025.07.07.25331048

**Authors:** Kelsie Raspin, Daisy Nowakowski, Joanne L Dickinson, Jessica Roydhouse

## Abstract

Advances in genomic technologies have driven a substantial shift in cancer care. Understanding patients’ experience of care and their associated outcomes is essential to effectively delivering precision care. These outcomes are usually evaluated through patient reported measures (PRMs), rather than administrative data. We conducted a systematic review of the literature to identify, describe and summarise the PRMs employed when patients with cancer underwent genetic testing. Search terms included cascade screening OR carrier screening OR genetic testing OR genetic counselling AND cancer AND patient reported outcomes AND NOT Review (Publication Type). A total of 474 articles were identified and following full text review, 46 studies were included for data extraction. We identified 98 measures, with 32 of them being genetics specific PRMs. These PRMs focussed on knowledge of genetics, perceptions, concerns, interest and acceptability. Majority of the included studies were conducted in the United States (n=27; 58.7%) and included breast cancer patients (n_breast_=14; n_breast&ovarian_=13). Given differences in legislative frameworks regarding the use of genetic information across countries, most included studies were conducted in countries where discrimination based on genetic features is illegal. It was also evident that studies in cancers other than breast are warranted and could inform standardised collection of key outcomes across the cancer spectrum. This review has identified, described and summarised what types of measures have been used when patients with cancer have undergone genetic testing and has highlighted the urgent need for development of additional PRMs in this area, particularly in cancers other than breast.

**Summary of Research Article:** We identified, described and summarised the measures published studies have used to gather a patients’ experience and outcomes related to genetic testing for their cancer. Our review identified 46 studies that used 98 different measures, 32 of which were associated with genetics specific topics, such as knowledge of genetics, perceptions, concerns, interest and acceptability of genetic testing. This review has highlighted that most studies have been conducted in the United States and included mainly breast cancer or breast and ovarian cancer patients.

## 1. INTRODUCTION

Genomic medicine has grown rapidly in recent years due to advances in genomic technologies and subsequent genetic discoveries which has enabled the delivery of precision care for those diagnosed with a range of cancers. The use of genetics in clinical care pathways has transformed screening, diagnosis, and treatment options for several common cancers, resulting in earlier diagnosis and better survival for patients with high-risk or metastatic disease (reviewed in Berger & Mardis, 2018 [1]). However, the integration of genomics in to cancer healthcare varies widely from country to country, and even within the same country due to inequitable access to genetic testing, perhaps in part due to a lack of resources and geographical remoteness [2].

A shift towards a more patient-centred healthcare system goes hand in hand with consumer/patient engagement and is paramount to the use of genetic information and effective delivery of precision medicine. Such initiatives, including one undertaken by Giri and colleagues (2022), highlight an important need for determining patient-reported outcome measures (PROMs) among men undergoing genetic testing for their cancer [3]. Genetic testing may have a psychosocial impact on patients, particularly for individuals at higher risk, such as those with a family history of disease [4]. Psychosocial outcomes are difficult to capture with administrative or clinical data and are best captured using PROMs. PROMs are a measurement based on a report that comes directly from the patient about their own health condition without amendment or interpretation from others [5]. PROMs are increasingly used in oncology, where their inclusion as part of routine cancer care has been shown to have a positive effect [6], including improved patient survival [7]. Patient reports of their experience are typically measured with patient reported experience measures (PREMs). PROMs and PREMs can be broadly described as patient-reported measures (PRMs). PRMs provide an invaluable insight into patient experiences, both as part of the research process and in clinical care as a patients’ perception of their disease symptoms, treatment experience and outcomes often differ from that of their clinicians [8].

Genomics has become widely integrated into the cancer healthcare setting and it is vital that the outcomes that matter most to patients continue to be captured, thus ensuring the delivery of patient-centred care and the successful translation of genomics into every day clinical practice. A critical first step is understanding what measures are being used, to inform future implementation. Thus, we have performed a systematic review of the literature surrounding the PRMs utilised when genetic testing for cancer has been undertaken. We aimed to identify, describe and summarise the PRMs used in these studies. PRM evaluation and recommendations for their use were not within the scope of this review.

## 2. METHODS

### 2.1 Search strategy

We conducted a PubMed literature search focusing on PRMs employed when patients with cancer underwent genetic testing. In the first instance, a series of pilot searches were conducted in March 2023 to identify relevant search terms. The following search criteria were used to identify the relevant studies; cascade screening OR carrier screening OR genetic testing OR genetic counselling AND cancer AND patient reported outcomes AND NOT Review (Publication Type). We conducted a pilot training exercise in which a reviewer (DN) screened the first 50 titles and abstracts to ensure that our search criteria were capturing the studies we expected. The search was performed without date restrictions but was limited to full-text articles, English-only articles and studies with an adult population over 18 years of age. The final search was conducted on the 2^nd^ of May 2025 and all articles were then imported into Covidence for review (Covidence systematic review software, Veritas Health Innovation, Melbourne, Australia. Available at www.covidence.org).

### 2.2 Eligibility criteria

Studies were only included where the outcome measure was reported by the patient, who was in receipt of a germline genetic test specifically for their cancer. Validation studies, protocols, meta-analyses, case reports, editorial comments and letters were excluded. Studies that only assessed clinical/demographic outcomes and that did not include solid cancers were also excluded.

### 2.3 Study selection and screening

Studies identified through the above search strategy were assessed for inclusion, initially by title and abstract. One reviewer assessed each title and abstract independently (DN), and a second reviewer (KR) assessed only the studies that were removed to determine if their exclusion was appropriate. Those articles that made it to full text review were screened against the eligibility criteria. Review decisions and key data were recorded using Covidence. When articles were excluded, reviewers (KR & DN) selected an exclusion reason from a drop-down list. Figure 1 shows a PRISMA flow diagram of the study selection and screening.

**Figure 1:**
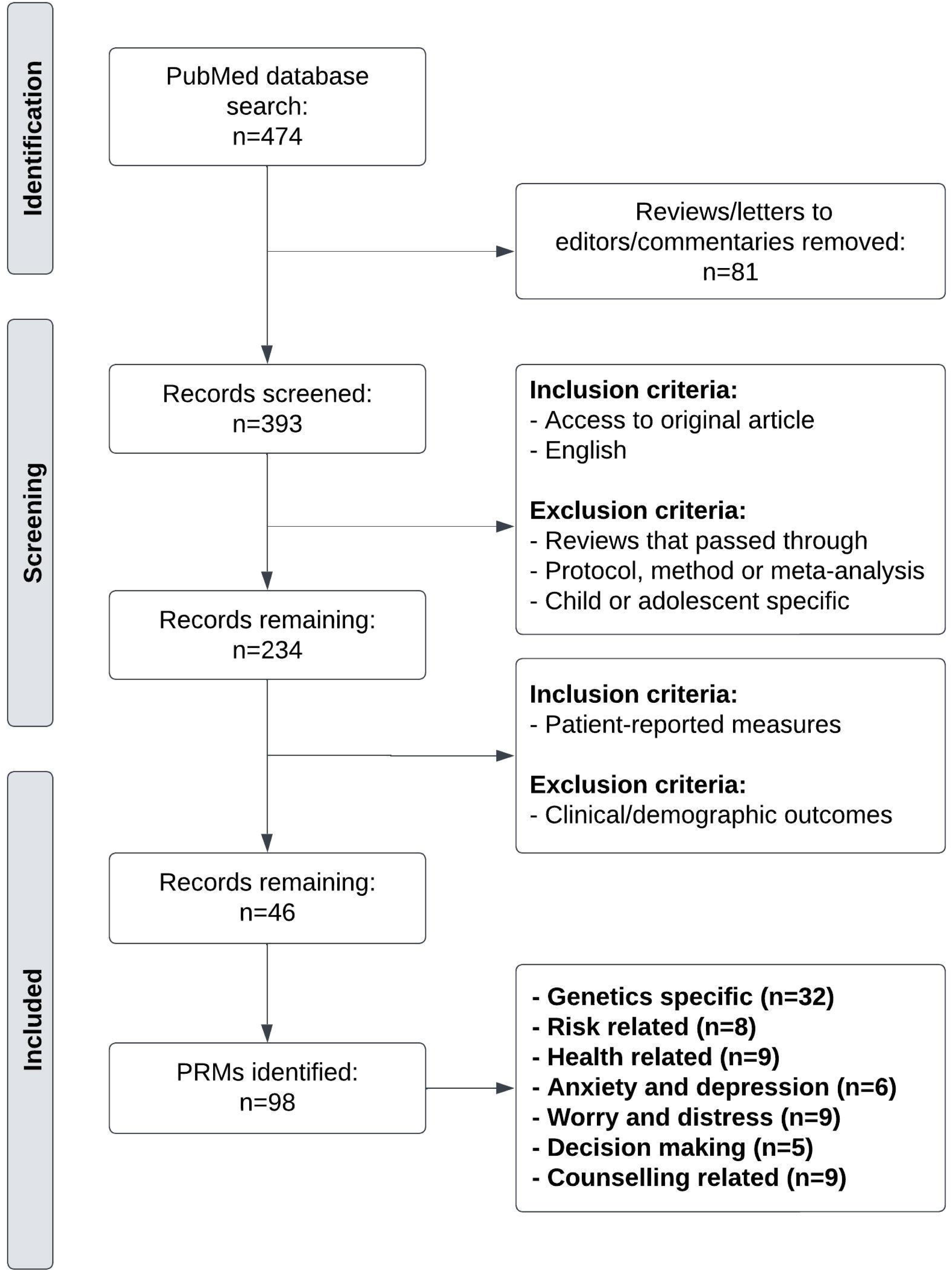
PRISMA flow diagram of the literature search in PubMed, conducted in May 2025.

### 2.4 Full text data extraction

Key information about each article was recorded in Covidence by one reviewer (KR), and checked by a second, independent reviewer (JR). The reviewer extracted data on the following areas: (1) domain/measure/tool title (measure), (2) domain/measure/tool acronym, (3) information about tool, (4) original development paper and adaptation, (5) article type, (6) year of publication, (7) country of study; as determined by description of study cohort, (8) study design, (9) length of study, (10) number of participants, and (11) type of cancer.

### 2.5 Classification and summary of evidence of PRMs

To better understand the PRMs used, the evidence supporting their use was reviewed. Citations, if any, for the measures provided in the studies were checked and the included measures were briefly described. We also noted if there had been any apparent adaptation of the measures for the relevant studies, and the evaluation, if any, of these adaptations. Measures were then grouped by their area of focus, (1) genetics specific PRMs, (2) risk related PRMs, (3) health related PRMs, (4) anxiety and depression PRMs, (5) worry and distress PRMs, (6) decision making PRMs, (7) genetic counselling related PRMs, and (8) other PRMs that do not fit into any other classification. Grouping was undertaken by one reviewer (KR) and checked by a second reviewer (JR).

## 3. RESULTS

The search identified 474 possible articles, from which 393 abstracts and titles were screened. Of these, 234 articles were deemed eligible following the aforementioned inclusion criteria and reviewed in full, of which, 46 were included for data extraction.

### 3.1 Summary and characteristics of the included studies

Of the 46 studies selected for inclusion, the majority were performed in the last decade (80.9%) and in the United States (58.7%), with more than half (54.3%), cohort studies. Altogether, 23.9% of studies were 2 years in length and 60.9% included <300 participants. Study designs ranged from randomised controlled trials (n=10) to the clinical setting (n=34). In total, 14 of the 46 studies included breast cancer patients, 13 included both breast and ovarian cancer patients and 9 included more than three cancer types. There were few studies including just colorectal (n=2), ovarian (n=4), and prostate cancer (n=4). A full summary of characteristics is shown in Table 1.

**Table 1.** Summary characteristics of studies included in the systematic review (n=46).

| Characteristics | Number of studies | % of studies |
| --- | --- | --- |
| Year of Publication |  |  |
| 2000-2005 | 1 | 2.2 |
| 2006-2010 | 3 | 6.5 |
| 2011-2015 | 8 | 17.4 |
| 2016-2020 | 15 | 32.6 |
| 2021-2025 | 19 | 41.3 |
| Country |  |  |
| Australia | 2 | 4.3 |
| Austria | 1 | 2.2 |
| Canada | 6 | 13.0 |
| France | 1 | 2.2 |
| Germany | 2 | 4.3 |
| Malaysia | 1 | 2.2 |
| Netherlands | 4 | 8.7 |
| Norway | 1 | 2.2 |
| United Kingdom | 1 | 2.2 |
| United States of America | 27 | 58.7 |
| Study Design |  |  |
| Case-Control | 2 | 4.3 |
| Cohort | 25 | 54.3 |
| Cross-Sectional | 2 | 4.3 |
| Non-randomised Experimental | 5 | 10.9 |
| Qualitative Research | 2 | 4.3 |
| Randomised Controlled Trial | 10 | 21.7 |
| Study Length (years) |  |  |
| 1 | 5 | 10.9 |
| 2 | 11 | 23.9 |
| 3 | 8 | 17.4 |
| 4 | 5 | 10.9 |
| 5 | 3 | 6.5 |
| 6 | 5 | 10.9 |
| 8 | 2 | 4.3 |
| Not reported | 7 | 15.2 |
| Number of Participants |  |  |
| 1-100 | 10 | 21.7 |
| 101-200 | 8 | 17.4 |
| 201-300 | 10 | 21.7 |
| 301-400 | 4 | 8.7 |
| 401-500 | 1 | 2.2 |
| 501-1000 | 7 | 15.2 |
| >1000 | 6 | 13.0 |
| Cancer Type |  |  |
| Breast | 14 | 30.4 |
| Breast and Ovarian | 13 | 28.3 |
| Colorectal | 2 | 4.3 |
| Ovarian | 4 | 8.7 |
| Prostate | 4 | 8.7 |
| >3 Cancer Types | 9 | 19.6 |

### 3.2 Categories of measures

We identified 98 measures used in the 46 included studies, which were grouped into eight different categories of classification. Of the included studies, the mean number of PRMs assessed in each study was between 4 and 5 (SD=2.24).

#### 3.2.1 Genetics specific PRMs

We identified 32 genetics specific PRMs (Table 2) which focussed on knowledge of genetics (n=8), perception of information (n=1), perceived utility of genetic testing (n=1), perceived personal control (n=1), concerns regarding genetic testing (n=1), interest in genetic testing (n=2), genetic testing experience and satisfaction (n=4), acceptability of genetic testing (n=2), genetic testing results (n=8) and risk management (n=4). Knowledge of genetics PRMs encompassed previously used and validated measures such as the Knowledge Scale [9], the Breast Cancer Genetic Counselling Knowledge Scale [10], the KnowGene Scale [11] and the ClinSeq Knowledge Scale [12–15], as well as study-specific measures used without apparent validation (n=3 studies). Genetic testing experience and satisfaction focussed on satisfaction with clinical services [16], experience and understanding of genetic testing (n=3 studies), reasons for declining [17], and perceived efficacy of screening [18]. Issues related to genetic testing results were mainly captured using the Multidimensional Impact of Cancer Risk Assessment (MICRA; n=19 studies). Nine others studies assessed a range of other genetic testing results PRMs, such as perception of process of results being returned, feelings about results, feelings about sharing results [19] and disclosure of genetic testing results [20,21]. Risk management PRMs were all study-specific measures that appeared to be used without validation. Measures assessed beliefs and attitudes about preventing cancer [22], behavioural intentions regarding cancer prevention [15,23], cancer prevention/screening behaviour decisions [17,24–27] and lifestyle changes in response to testing results [28].

**Table 2:**
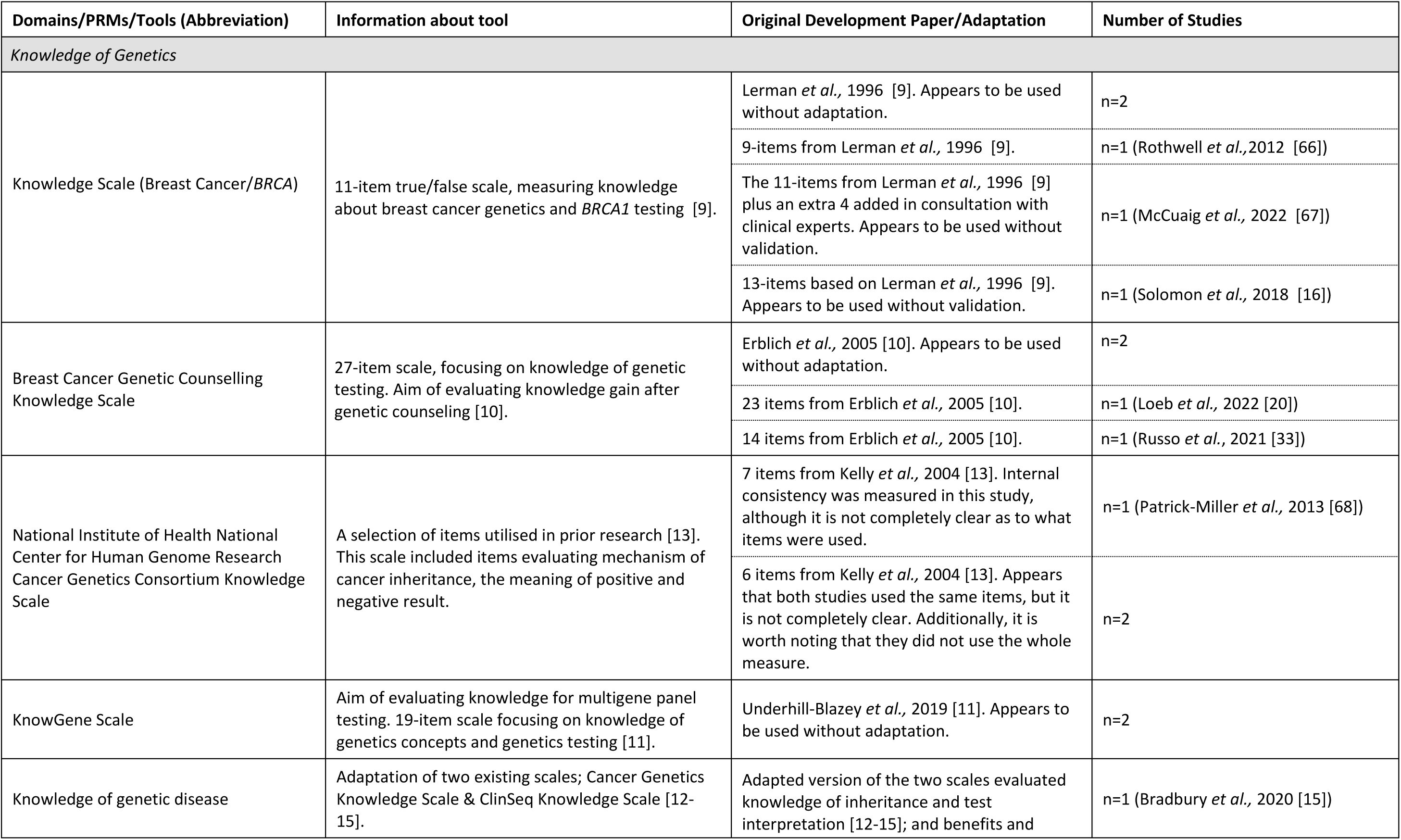
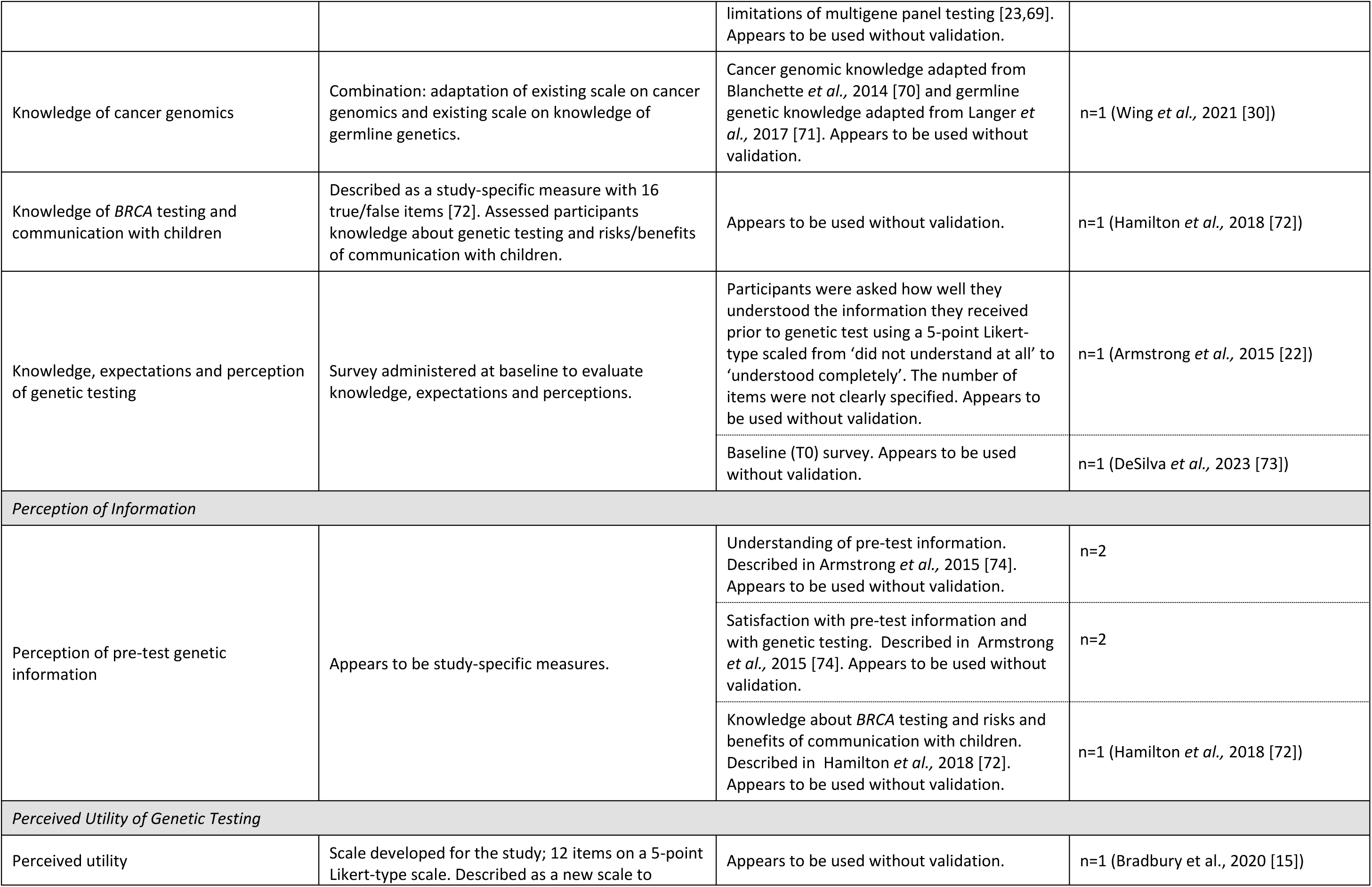
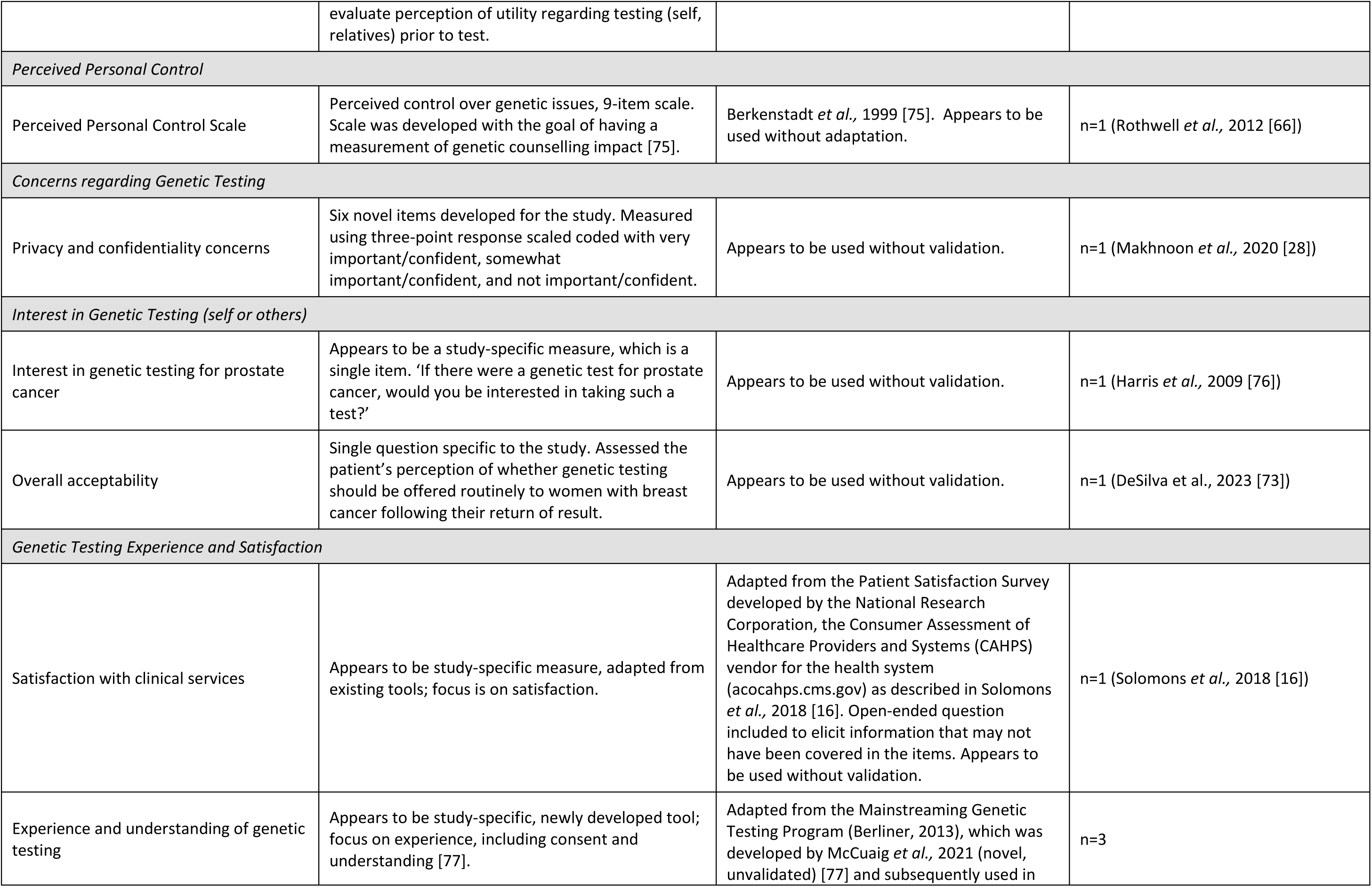
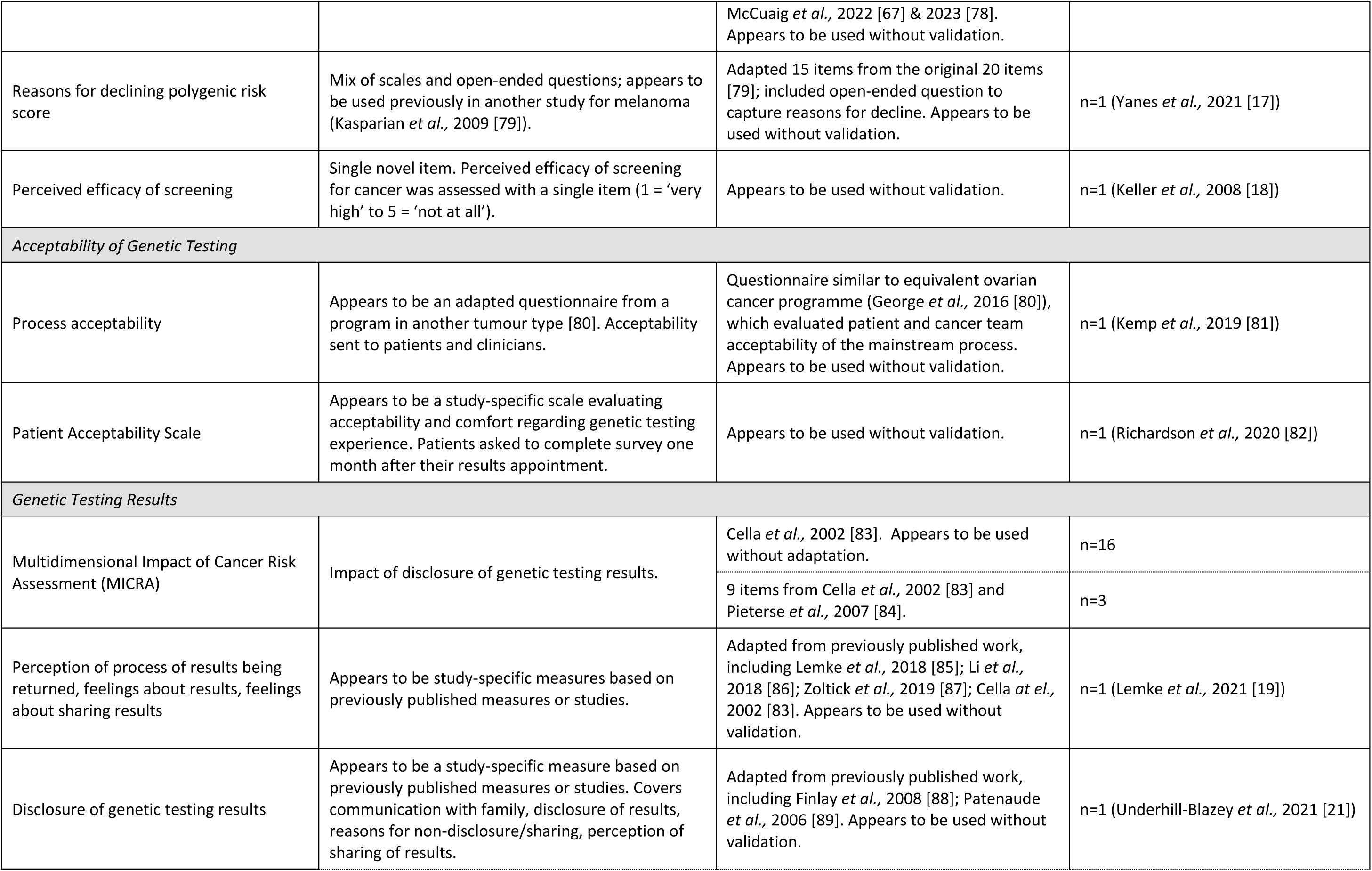
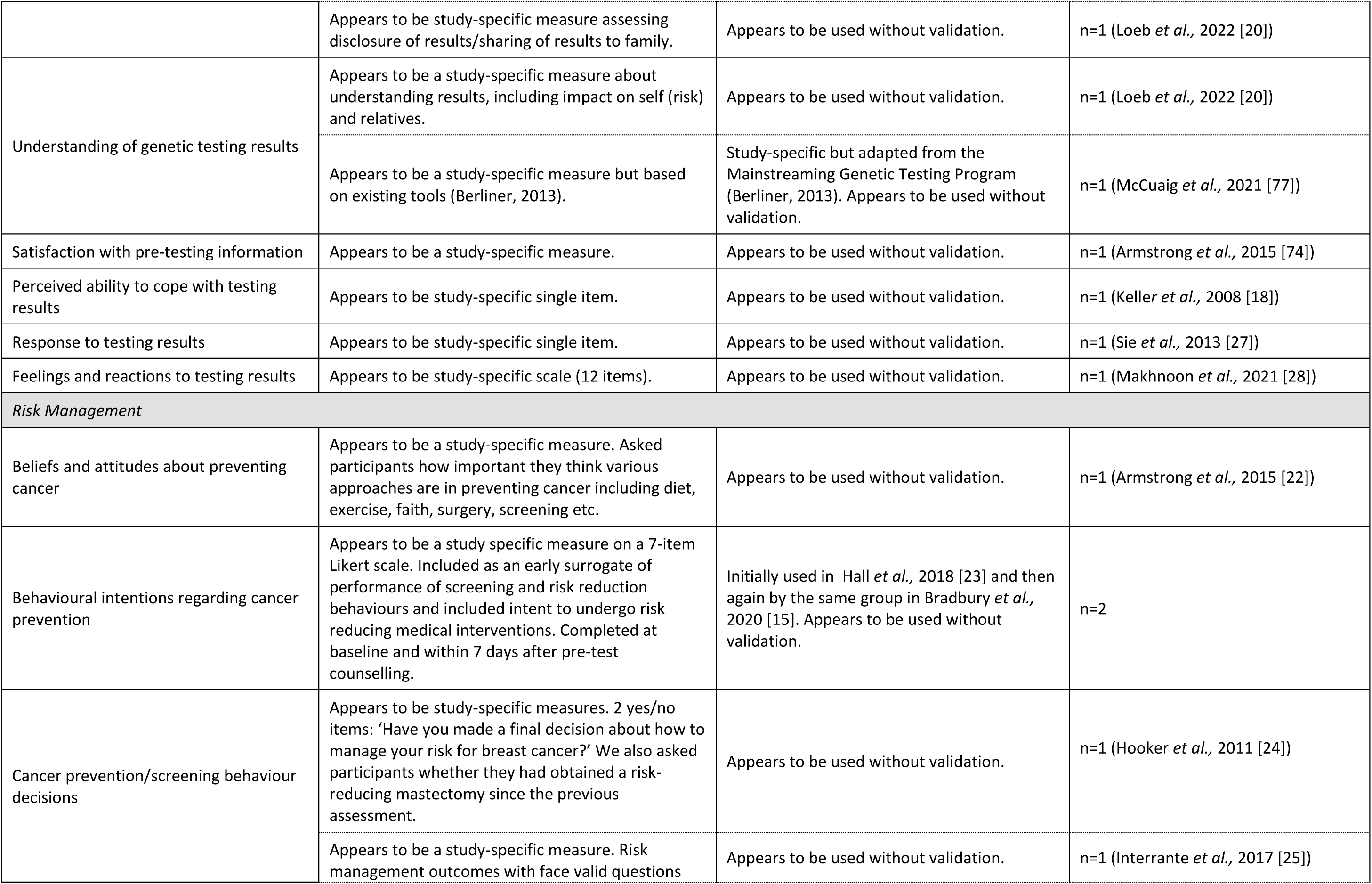
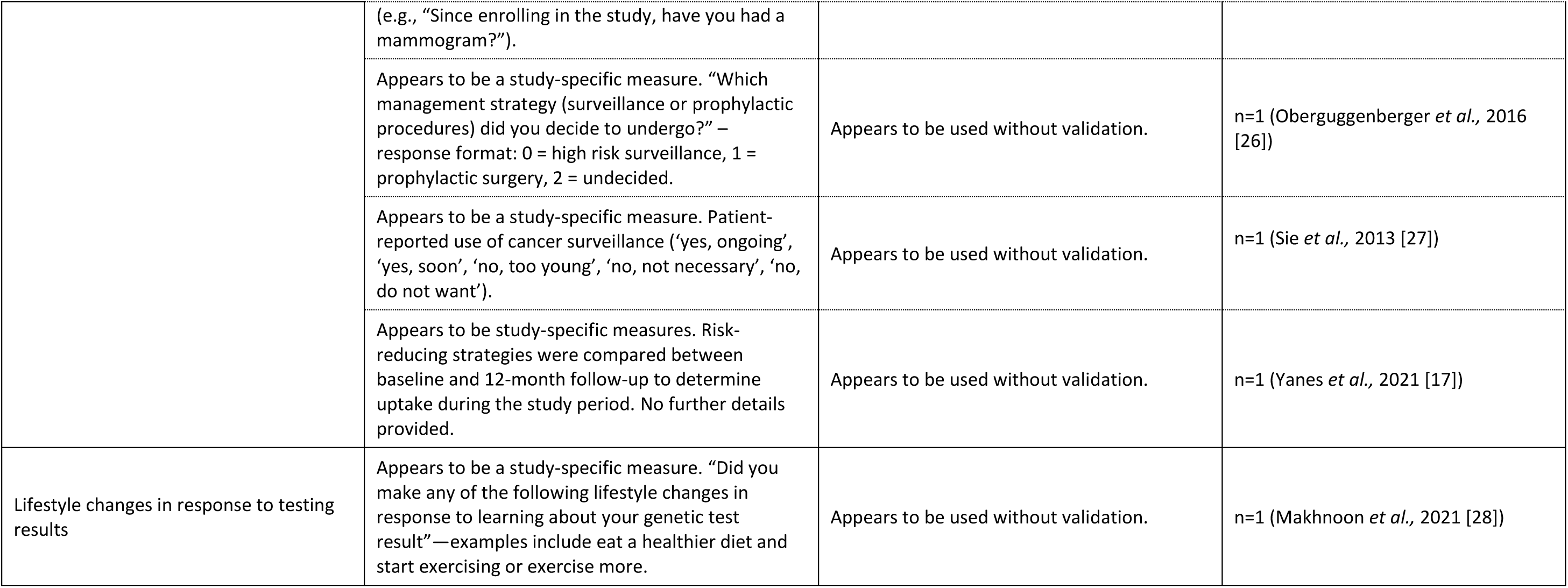
Overview of the genetics specific PRMs identified by the systematic literature search.

Other genetics specific measures that we identified but did not classify as a PRM were family history of cancer and genetic test result recall. Four studies asked participants whether they had a family history of cancer, yes/no. Eijzenga *et al.,* 2015 [29] asked the participants whether they had a history of genetic counselling for cancer in their family. In terms of result recall, two studies asked participants to recall their genetic test result which was compared to the result recorded in their medical record [22,30].

#### 3.2.2 Risk related PRMs

Eight risk related PRMs were ustilised in 10 different studies (Table 3). PRMs assessed numeracy knowledge (n=2), health literacy (n=2), perceived cancer risk (n=3) and received recurrence risk (n=1). Numeracy knowledge measures were used to determine the effects of low numeracy on comprehension of risk communications and utility elicitations [31–33]. The most commonly used risk related PRM was perception of cancer risk(s), which was included in five studies. Studies used a variety of formats but all, except one [34], were a study-specific measure used without validation.

**Table 3.** Overview of the risk related PRMs identified by the systematic literature search.

| Domains/PRMs/Tools (Abbreviation) | Information about tool | Original Development Paper/Adaptation | Number of Studies |
| --- | --- | --- | --- |
| *Numeracy Knowledge* |  |  |  |
| Numeracy scale | Three item scale evaluating understanding of probability and other mathematical concepts; appears to be study-specific [90]. | Schwartz *et al.*, 1997 [90]. Appears to be used without adaptation. | n=2 |
| Subjective Numeracy Scale | Evaluation of understanding of risk interpretation, including for health economic applications (e.g., utility or trade-off); 8-item scale [91,92]. | Fagerlin *et al.*, 2007 [91]. Appears to be used without adaptation. | n=1 (Kukafka *et al.*, 2022 [31]) |
| *Health Literacy* |  |  |  |
| eHealth Literacy Scale (eHEALS) | Health literacy scale, including understanding of computer/eHealth. 8-item scale [93]. | Norman *et al.*, 2006 [93]. Appears to be used without adaptation. | n=1 (Kukafka *et al.*, 2022 [31]) |
| Three-item screening questionnaire for health literacy | Brief 3-item screening questionnaire evaluating health literacy [94]. | Study used three items from Chew *et al.*, 2004 [94]. Appears to be used without adaptation. | n=1 (Russo *et al.*, 2021 [33]) |
| *Perceived Cancer Risk* |  |  |  |
| Perception of cancer risk(s) | Appears to be a study-specific measure. Assessed using measures of quantitative (chance from 0-100%) and comparative (Likert 5-point scale) perceived risk. Accuracy of perceived risks was compared to those calculated risks based on validated models that incorporate personal/family cancer history (e.g. Gail [95,96]) and test results [97]. | Appears to be used without validation. | n=1 (Armstrong *et al.*, 2015 [22]) |
|  | Two Likert-style items previously employed [34] | Audrain *et al.*, 1997 [34]. Appears to be used without adaptation. | n=1 (Schwartz *et al.*, 2003 [98]) |
|  | Appears to be a study-specific measure. Evaluated using a single ranking question (0-100%). | Appears to be used without validation. | n=1 (McCuaig *et al.*, 2021 [77]) |
|  | Visual scale, specific to the study. A visual scale of 0-100 was used to measure lifetime risk perception of cancer occurrence. | Appears to be used without validation. | n=1 (Sie *et al.*, 2013 [27]) |
|  | Study-specific survey asking women whether their own perception of cancer risk changed after their genetic test. | Appears to be used without validation. | n=1 (Morman *et al.*, 2017 [99]) |
| Perception of vulnerability to and lifetime risk of cancer | Appears to be a study-specific measure, evaluating perceived risk for self and family, as well as the perceived threat of cancer. Threat of cancer was marked on a 100mm visual analogue scale (0 = 'not at all' to 100 = 'very threatening'). | Perceived vulnerability and risk to family may have been adapted from Codori *et al.*, 1999 [100]. | n=1 (Keller *et al.*, 2008 [18]) |
| Distress due to Hereditary Disorder (DHD) Scale | Study specific 3-item scale. Concerns about their relatives' cancer risk, their own cancer risk and the degree to which these worries interfere with daily functioning. | Modified version of the Cancer Worry Scale - Lerman *et al.*, 1991 [37]. | n=1 (Keller *et al.*, 2008 [18]) |
| *Perceived Recurrence Risk* |  |  |  |
| Perception of risk of breast cancer recurrence | Appears to be a study-specific single item. "What do you think the chance is that your breast cancer will come back or spread to other parts of the body? Please choose a number between 0 % (no chance) and 100 % (definitely will)." | Appears to be used without validation. | n=1 (Evans *et al.*, 2016 [101]) |

#### 3.2.3 Health related PRMs

Health related PRMs (Table 4) evaluated health status/quality of life (n=3), performance status (n=1), sexual health (n=2) and symptoms (not specific to sexual health; n=3). All PRMs that were utilised were previously developed and used without apparent adaptation. The most commonly used health related PRM was the SF-12 Health Survey (SF-12) which was used in four studies. The ECOG Performance Status, which is normally clinician-reported, was used as a PRM in Hamilton *et al.,* 2021 [35].

**Table 4.** Overview of the health related PRMs identified by the systematic literature search.

| Domains/PRMs/Tools (Abbreviation) | Information about tool | Original Development Paper/Adaptation | Number of Studies |
| --- | --- | --- | --- |
| *Health Status/Quality of Life* |  |  |  |
| SF-36 Health Survey (SF-36) | Generic multi-dimensional measure of health-related quality of life; used in many conditions, not specific to cancer [102]. | Ware & Sherbourne, 1992 [102]. Appears to be used without adaptation. | n=2 |
| SF-12 Health Survey (SF-12) | Generic multi-dimensional measure of health-related quality of life; used in many conditions, not specific to cancer [103]. | Ware *et al.*, 1994 [103]. Appears to be used without adaptation. | n=4 |
| EORTC Quality of Life Questionnaire – Core Questionnaire (EORTC-QLQ-C30) | Multi-dimensional measure of health-related quality of life. Specific to cancer but not a specific type of cancer [104]. | EORTC Quality of Life Group, 1995 [104]. Appears to be used without adaptation. | n=1 (Sie *et al.*, 2013 [27]) |
| *Performance Status* |  |  |  |
| Eastern Cooperative Oncology Group Performance Status (ECOG Performance Status) | Usually a clinician-reported measure in cancer, describing functional ability [105]. | Eastern Cooperative Oncology Group, 1982 [105]. Used as a patient-reported measure in the study. Appears to be used without validation. | n=1 (Hamilton *et al.*, 2021 [35]) |
| *Sexual Health* |  |  |  |
| Sexual Health Inventory for Men (SHIM) (or Erectile Function – 5 items; IEF-5) | Shortened version of the International Index of Erectile Function [59]. | Rosen *et al.*, 1999 [106]. Appears to be used without adaptation. | n=1 (Harper *et al.*, 2023 [54]) |
| American Urological Association (AUA) Symptom Scale | Scale to measure symptoms of benign prostatic hyperplasia; 7 items [107]. | Barry *et al.*, 1992 [107]. Appears to be used without adaptation. | n=1 (Harper *et al.*, 2023 [54]) |
| *Symptoms (not specific to sexual health)* |  |  |  |
| Symptom Checklist-90-Revised (SCL-90-R®) | Checklist focusing on psychological symptoms, not specific to cancer [108]. Short form of the SCL-90 [109]. | Derogatis *et al.*, 1973 [109]. Appears to be used without adaptation. | n=1 (Schwartz *et al.*, 2003 [98]) |
| Brief Symptom Inventory (BSI) | Self-reported symptom measure, focusing on psychological symptoms. Adapted from the SCL-90-R [108-110]. Not specific to cancer [111]. | Derogatis *et al.*, 1983 [110]. Appears to be used without adaptation. | n=2 |
| Gastrointestinal scale from the Giessen Complaints Inventory | Set of self-reported physical complaints, not specific to cancer. 7-item scale (range 0-24). Used to look at “somatic expression of cancer fears as a potential response to cancer-related threat" [18]. | Braehler & Scheer 1995 [112]. Appears to be used without adaptation. | n=1 (Keller *et al.*, 2008 [18]) |

#### 3.2.4 Anxiety and depression PRMs

We identified six anxiety and depression PRMs (Table 5) which were all previously developed and used without apparent adaptation in the included studies. The Hospital Anxiety and Depression Scale (HADS) [36] was the most commonly used anxiety and depression PRM (n=13 studies), followed by the State-Trait Anxiety Inventory (STAI; n=7 studies).

**Table 5.** Overview of the anxiety and depression PRMs identified by the systematic literature search.

| **Domains/PRMs/Tools (Abbreviation)** | **Information about tool** | **Original Development Paper/Adaptation** | **Number of Studies** |
| --- | --- | --- | --- |
| Hospital Anxiety and Depression Scale (HADS) | Generic measure of anxiety and depression; not specific to cancer [36]. | Zigmond & Snaith, 1983 [36]. Appears to be used without adaptation. | n=13 |
| Depression Anxiety Stress Scales Short Form (DASS-21) | Shorter version of a longer generic instrument measuring depression, anxiety, and stress; not specific to cancer [113,114]. | Henry & Crawford, 2005 [115]. Appears to be used without adaptation. | n=1 (DeSilva *et al.*, 2023 [73]) |
| Memorial Anxiety Scale for Prostate Cancer (MAX-PC) | Measures anxiety in prostate cancer; 18 items, covers prostate cancer anxiety and PSA testing anxiety [116]. | Roth *et al.*, 2003 [117]. Appears to be used without adaptation. | n=1 (Harper *et al.*, 2023 [54]) |
| State-Trait Anxiety Inventory (STAI) | Generic measure of anxiety (state and trait) [118]; not specific to cancer [118]. | Speilberger *et al.*, 1983 [119]. Appears to be used without adaptation. | n=7 |
| Generalized Anxiety Disorder – 7 item (GAD-7) | Generic measure of anxiety disorder; 7 item scale [120]. | Spitzer *et al.*, 2006 [120]. Appears to be used without adaptation. | n=3 |
| Patient Health Questionnaire – 4 item (PHQ-4) | Generic measure of depression and anxiety; can be used for screening [121]. | Kroenke *et al.*, 2009 [121]. Appears to be used without adaptation. | n=1 (Solomons *et al.*, 2018 [16]) |

#### 3.2.5 Worry and distress PRMs

In terms of worry and distress, six PRMs assessed worry and uncertainty and three evaluated distress (Table 6). The Cancer Worry Scale (CWS) [37] was used most often to measure worry and uncertainty. The CWS was used to assess breast cancer-related worries in four studies and was further adapted without validation in one other [38]. The Impact of Events Scale – 15 items (IES-15) [39,40] was the most commonly used distress PRM. Thirteen studies used the IES-15 without apparent adaptation, and one used two of its items [18]. The remaining PRMs included four previously used and validated measures, as well as two study-specific measures.

**Table 6.**
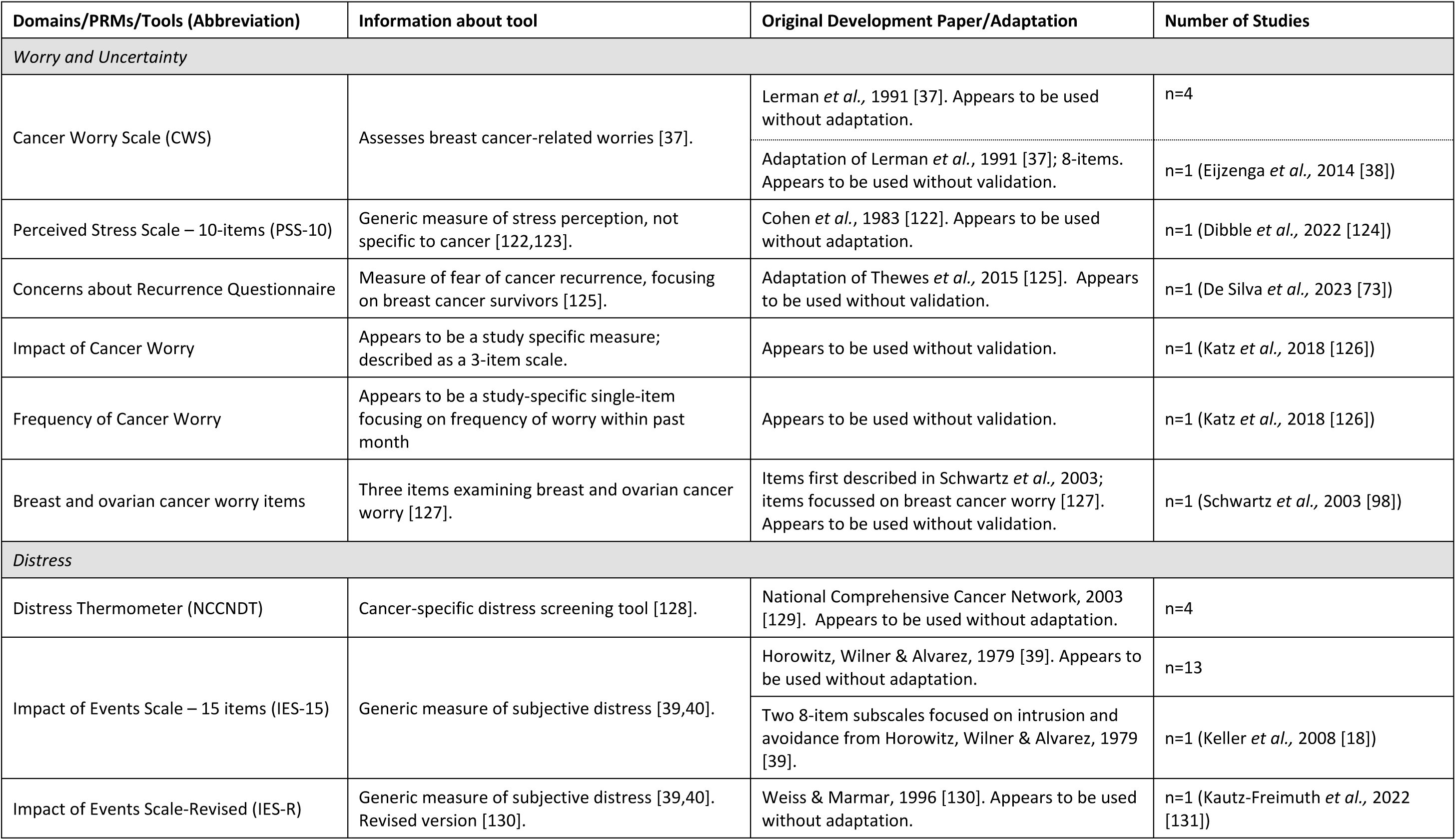
Overview of the worry and distress PRMs identified by the systematic literature search.

#### 3.2.6 Decision making PRMs

Decision making PRMs included five measures that were all previously developed and used in most of the included studies without apparent adaptation (Table 7). The PRMs assessed decisional conflict, satisfaction with decisions, decision self-efficacy, decision regret and control preferences. The most commonly used PRM was the Decisional Conflict Scale (DCS) [41] which measures the perception of decision-making, including uncertainty regarding decisions. Six studies utilised the DCS without apparent adaptation, and one study by Sie *et al.,* 2013 [27] removed one item from the DCS as the authors determined that the item was not relevant to genetic testing. We did not find evidence of validation or testing of this modified instrument.

**Table 7.** Overview of the decision making PRMs identified by the systematic literature search.

| Domains/PRMs/Tools (Abbreviation) | Information about tool | Original Development Paper/Adaptation | Number of Studies |
| --- | --- | --- | --- |
| Decisional Conflict Scale (DCS) | Perception of decision-making, including uncertainty regarding decisions [41]. | O'Connor, 1995 [41]. Appears to be used without adaptation. | n=6 |
|  |  | Adaptation of O'Connor, 1995 [41]; 1 item removed as seen as not relevant for genetic testing. Appears to be used without validation. | n=1 (Sie *et al.*, 2013 [27]) |
| Satisfaction with Decision Scale (SWD) | Satisfaction with care decisions [132]. | Holmes-Rovner *et al.*, 1996 [132]. Appears to be used without adaptation. | n=3 |
| Decision Self Efficacy Scale (DSES) | Self-confidence or belief in one's ability to make decisions [133]. | Cranney *et al.*, 2002 [134]. Appears to be used without adaptation. | n=2 |
| Decision Regret Scale | Measures regret after healthcare decision-making [135]. | Brehaut *et al.*, 2003 [135]. Appears to be used without adaptation. | n=2 |
| Control Preferences Scale | Measures treatment decision-making preferences [136]. | Degner, Sloan & Venkatesh, 1997 [136]. Appears to be used without adaptation. | n=1 (Kukafka *et al.*, 2022 [31]) |

#### 3.2.7 Genetic counselling related PRMs

We identified nine genetic counselling related PRMs (Table 8) which focussed on genetic counselling satisfaction (n=3), outcomes (n=1), worry (n=1), self-efficacy (n=1), perceptions (n=1) and preferences (n=2). The most commonly used PRM was the Genetic Counselling Satisfaction Scale (GCSS; n=6) [42].

**Table 8.**
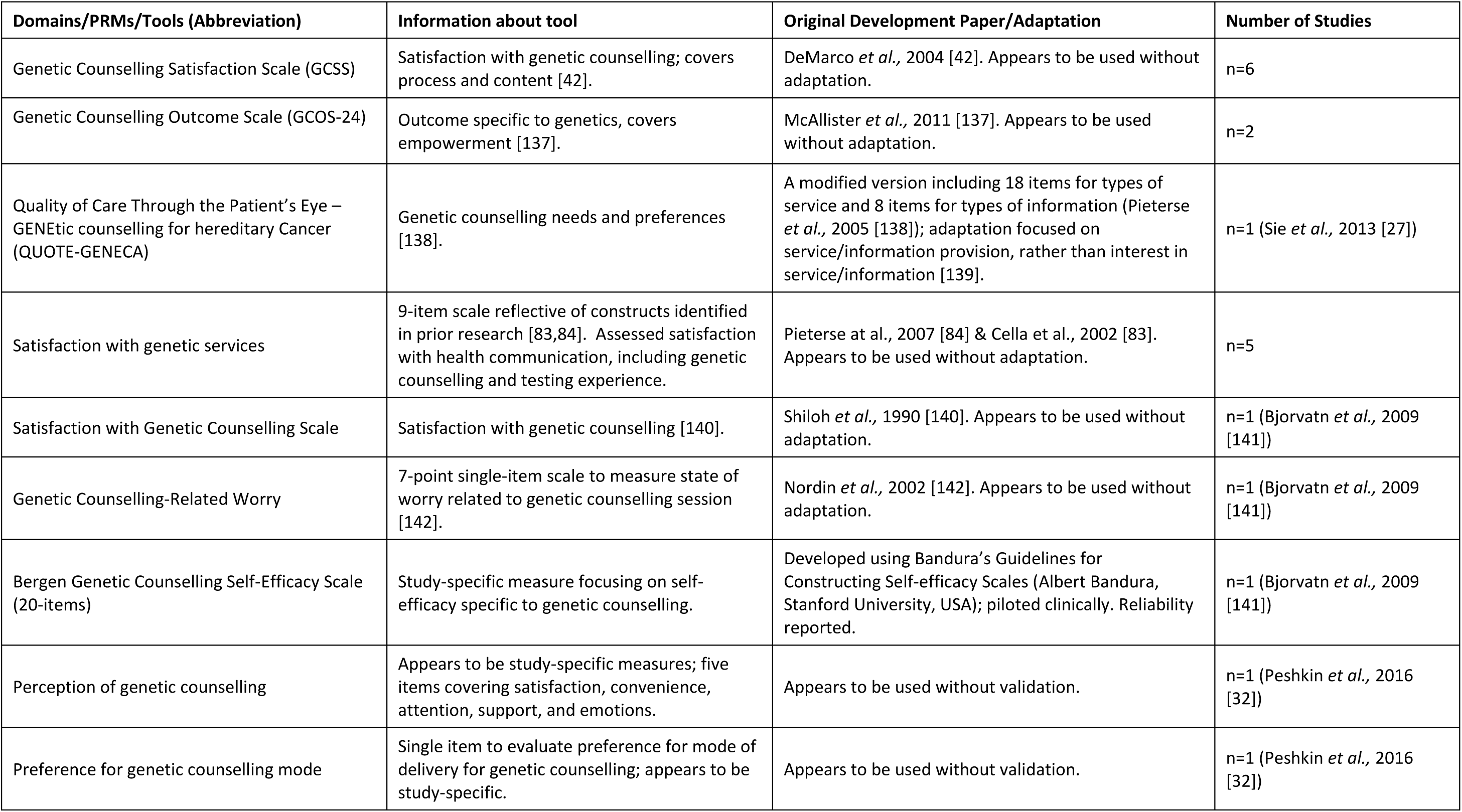
Overview of the counselling related PRMs identified by the systematic literature search.

#### 3.2.8 Other PRMs

PRMs not covered under the above categories (Supplementary Table 1) focussed on perceptions of surgery outcomes (n=1) and chemotherapy (n=1), telemedicine (n=3), medical threat (n=1), perceived social support (n=1), ease of access to services (n=1), dispositional optimism (n=1) and quality of parent-child communication relationship (n=1). The telemedicine PRMs measured perception and experience, as well as satisfaction and acceptability of telephone counselling sessions.

## 4. DISCUSSION

This review is the first, to our knowledge, to identify, describe and summarise the PRMs used when genetic testing for cancer has been undertaken across the research and clinical spectrum. Across the 46 studies included in our review, 98 PRMs were identified, spanning the following categories: genetics specific, risk related, health related, anxiety and depression, worry and distress, decision making and counselling related. The most common category was genetics specific with 32 PRMs; knowledge of genetics and genetic testing results were the most commonly measured in this category.

Approximately 80% of the included studies were published in the last 10 years, which is consistent with the significant genetic discoveries for cancer made within this period (recently reviewed in Hall *et al.,* 2024 [43]). This is also consistent with the growing push towards more patient-centred care and increased use of PRMs in the clinical management of cancer, for example, PRO collection in Phase 1 cancer trials has been increasing over time [44]. Genetic testing has the potential to reduce cancer morbidity and mortality as genetic information can inform disease management, including eligibility for gene-based targeted therapies and/or clinical trials, as well as early screening strategies of at-risk individuals (i.e., those with a family history). Our review also had overrepresentation of studies conducted in the United States (58.7%). Discrimination on the basis of genetic features is illegal in the United States through the Genetic Information Nondiscrimination Act of 2008 (GINA) [45]. It is also illegal in the European Union (through the Charter of Fundamental Rights of the European Union) [46], whereas in other countries, such as Australia, there is currently no such protection for those wishing to undertake genetic testing. However, in September 2024, the Australian Government announced that it would introduce a total ban on the use of genetic test results in life insurance underwriting, though legislation is yet to be enacted. It is expected that this will have a positive impact on the uptake of genetic testing in Australia, which may see more Australian studies focussed on PRM collection in genetic testing for cancer over the coming decade. Furthermore, such differences in legislation pertaining to genetic testing may have influenced studies in this area and indeed may have led to very different patient perceptions and outcomes in different countries [47–50]. This highlights that the legislative frameworks shape engagement of the public in health initiatives and is worthy of investigation in different jurisdictions. Lastly, there was no apparent difference in the types of PRMs utilised between the different study designs, for example, randomised controlled trials (n=10) *versus* a clinical setting (n=34), although formal analyses were not undertaken.

The majority of studies that met our inclusion criteria included patients with breast cancer, or breast and ovarian cancer (n=27; 58.7%). This is not surprising, as genetic research into breast and ovarian cancer has led the way in revealing the risk genes conferring solid cancers and this knowledge has underpinned genomic medicine for these cancers, including screening, diagnosis, and the development of precision treatment strategies [51]. Specifically, genetic testing in multiple solid cancers, particularly breast and ovarian cancer, is informing patient eligibility for targeted therapies (such as poly-ADP-ribose polymerase inhibitors (iPARP) in *BRCA2* variant positive tumours) and/or clinical trials and is readily offered in Western countries. Whereas, despite having the highest genetic component of any common cancer (∼58% [52] *vs* 31% in breast [53]), there is a lack of known high-risk genes for prostate cancer, which has hindered the implementation of genetic testing in the clinical space. This explains why we identified very few studies exploring the PRMs regarding genetic testing for prostate cancer (n=4; 8.7%). The types of PRMs used by the prostate cancer studies were similar to the breast cancer studies, except for the study conducted by Harper and colleagues (2023) [54], whose PRMs focussed on sexual health (SHIM), symptoms (AUASS) and prostate cancer related anxiety (MAX-PC). The two studies on colorectal cancer also included similar PRMs to the remaining studies [18,28]. As genetic testing continues to grow in oncology, additional studies in other cancer types, such as prostate and colorectal cancer, would be beneficial and could inform standardised collection of key outcomes across the cancer spectrum.

Previous reviews in this area are few - only two studies have reviewed PROs following genetic testing for cancer [55,56]. The most recent and relevant by Kalra and colleagues (2025) [56] assessed the uptake and PROs of germline genetic testing for cancer associated genes at or after a cancer diagnosis by a non-genetically trained member of the healthcare team (termed ‘mainstreaming’). They concluded that mainstreaming genetic testing uptake is associated with high uptake and satisfaction and low decisional conflict, regret and post-test distress [56]. This systematic review and meta-analysis included 29 studies published up until June 2024, of which, only seven studies overlapped with this current systematic review. The authors excluded studies that 1) offered genetic counselling by a clinical geneticist or genetic counsellor (either wholly or in part), 2) reported only qualitative research, and 3) exclusively offered cascade testing [56]. These were included in our review to ensure that we captured a broad representation of patients undergoing genetic testing. For example, the Kalra *et al.,* (2025) [56] published search and inclusion criteria effectively excluded studies in colorectal cancer. Further, there was no discussion of counselling related PRMs, in addition to genetics specific, risk related and health related PRMs. In comparison, our review did not seek to quantify the uptake of genetic testing or any associations with PRMs, instead we sought to identify, describe and broadly summarise all the PRMs currently used in any genetic testing for cancer setting. A second review focussed on breast and ovarian cancer, in conjunction with familial hypercholesterolemia and Lynch syndrome [55]. They identified 24 studies published between 1996 and 2021, reporting on 12 validated PROM instruments. All of these studies were captured by our search criteria however, only two met our inclusion criteria and were assessed in our review, namely Bradbury *et al.,* (2020) [15] and Underhill-Blazey *et al.,* (2021) [21]. Furthermore, Ringwald and colleagues (2016) described psychological distress, anxiety and depression of cancer affected *BRCA1/2* variant carriers across 1,243 studies [57], however they did not include patient reported in their search terms. Although psychological outcome measures (IES, MICRA and PSS), depression measures (HADS, Center of Epidemiologic Studies-Depression Scale, SCL-90, Hopkins Symptom Checklist, and Positive and Negative Affect Scale) and anxiety measures (HADS, STAI and Hopkins Symptom Checklist) were similar to the PRMs identified here, however none of the studies overlapped. Another systematic review identified studies that measured the perceived risk, psychological, and/or behavioural impacts of genetic testing on individuals, but the search was not limited to cancer or a specific disease [58]. They suggested that genetic testing for various disorders does not seem to lead to adverse consequences for carriers or non-carriers. A study by Menko and colleagues (2019) explored studies where *BRCA1/2* variant carriers informed at-risk family members about the option for presymptomatic DNA testing [59]. They found that the uptake of presymptomatic testing is disappointing. Two other reviews within the last 10 years were also identified, however they did not focus on PRMs following receipt of a genetic result instead they reported on psychosocial, behavioural and clinical outcomes which were not reported by the patient [60,61]. Although such studies can provide information and insight into general outcomes following genetic testing for cancer, PRMs directly measure the patients journey and provide valuable insight into patient preferences and opinions, therefore are much more informative [62].

One challenge identified in our review is that not all studies used PRMs that had evidence of validation as part of their development; some studies appeared to use validated instruments with adaptations without evidence of testing of these adaptations, and others used instruments that seemed study-specific and had no clear evidence of validation. The validity of a measure is an essential component in considering its use. For example, content validity, or the extent to which the measure covers the concept or construct of interest [63], is considered a core measurement property to the extent that it should be evaluated before other measurement properties such as reliability [64]. Future studies aiming to incorporate the patient voice via PRMs should prioritise the use of measures with evidence of validity. If study-specific or adapted measures were employed due to the lack of validated measures covering concepts of interest, this suggests a need for the development of additional measures in this area.

There are several strengthens and limitations of our systematic review. In terms of strengths, we developed a comprehensive search strategy following a series of pilot searches and we undertook quality checks at each stage to ensure we did not miss any study of relevance. Two independent reviewers also double checked the inclusion of studies and reviewed the identified PRMs. Most notably, we carefully examined each PRM, including the original citation to assess whether it was validated, followed by determining whether it was adapted in the included study. This was also double checked by a second reviewer. We must also acknowledge our limitations. Firstly, our literature search was conducted in only one database, PubMed. However, PubMed is the preferred search engine for this area of research and is the largest database for medical research. Secondly, we did not include articles in languages other than English; yet, evidence suggests that exclusion of non-English studies is unlikely to affect results [65]. We also included studies only in adult participants, which means that our results are valid only for adults who have been diagnosed with cancer. Lastly, though we did provide some description of the measures (i.e., ad hoc or not), a full COSMIN review of measurement properties was outside of the scope of this work. Measurement properties should be considered prior to selecting an instrument.

Our review provides valuable evidence regarding the PRMs used when genetic testing for cancer has previously been offered in the research and clinical setting. It has highlighted the lack of validity of PRMs used by such studies, however it is not clear whether the use of unvalidated PRMs was due to a lack of validated measures covering concepts of interest. If it is, then there is an urgent need for development of additional PRMs in this area. Our review also highlighted that there is a significant overrepresentation of studies in the breast and ovarian cancer setting. As genetic testing continues to grow in oncology, additional studies in other cancer types are warranted and could inform standardised collection of key outcomes. There is also a need for studies to be undertaken in a range of contexts given the influence of legislative frameworks regarding discrimination based on genetics. Furthermore, the studies included in this review are not only vital in determining what kinds of measures have been used in the genetic testing for cancer space but also help determine which outcomes are most important to patients. Understanding patients’ perceptions and expectations are pivotal to making sure genetic testing for cancer is patient centred. This information will enable the effective implementation and integration of genetic testing into clinical cancer care.

## Data Availability

All data is provided in the supplementary material.

## AUTHOR CONTRIBUTIONS

KR, JLD and JR conceived the review topic. DN performed the PubMed search, and KR and DN reviewed the titles and abstracts. KR and JR reviewed the full texts and recorded the key information. KR wrote the original draft and formatted the figures and tables. All authors reviewed and edited the manuscript.

## REFERENCES

1 Berger, M. F. & Mardis, E. R. The emerging clinical relevance of genomics in cancer medicine. Nat Rev Clin Oncol 15, 353–365 (2018). 10.1038/s41571-018-0002-6

2 Swami, N., Yamoah, K., Mahal, B. A. & Dee, E. C. The right to be screened: Identifying and addressing inequities in genetic screening. Lancet Reg Health Am 11, 100251 (2022). 10.1016/j.lana.2022.100251

3 Giri, V. N., Hartman, R. L., Gross, L. & Keith, S. W. Patient-reported outcomes among males undergoing prostate cancer germline testing: Interim results from the Prostate Cancer Genetic Risk, Experience, and Support Study (PROGRESS) Registry. Journal of Clinical Oncology 40 (2022).

4 Oliveri, S., Ferrari, F., Manfrinati, A. & Pravettoni, G. A Systematic Review of the Psychological Implications of Genetic Testing: A Comparative Analysis Among Cardiovascular, Neurodegenerative and Cancer Diseases. Front Genet 9, 624 (2018). 10.3389/fgene.2018.00624

5 Health, U. S. D. o., et al. Guidance for industry: patient-reported outcome measures: use in medical product development to support labeling claims: draft guidance. Health Qual Life Outcomes 4, 79 (2006). 10.1186/1477-7525-4-79

6 Yang, L. Y., Manhas, D. S., Howard, A. F. & Olson, R. A. Patient-reported outcome use in oncology: a systematic review of the impact on patient-clinician communication. Support Care Cancer 26, 41–60 (2018). 10.1007/s00520-017-3865-7

7 Basch, E. et al. Overall Survival Results of a Trial Assessing Patient-Reported Outcomes for Symptom Monitoring During Routine Cancer Treatment. Jama-J Am Med Assoc 318, 197–198 (2017). 10.1001/jama.2017.7156

8 Laugsand, E. A. et al. Health care providers underestimate symptom intensities of cancer patients: a multicenter European study. Health Qual Life Outcomes 8, 104 (2010). 10.1186/1477-7525-8-104

9 Lerman, C. et al. Controlled trial of pretest education approaches to enhance informed decision-making for BRCA1 gene testing. J Natl Cancer Inst 89, 148–157 (1997). 10.1093/jnci/89.2.148

10 Erblich, J. et al. Development and validation of a Breast Cancer Genetic Counseling Knowledge Questionnaire. Patient Educ Couns 56, 182–191 (2005). 10.1016/j.pec.2004.02.007

11 Underhill-Blazey, M. et al. Development and testing of the KnowGene scale to assess general cancer genetic knowledge related to multigene panel testing. Patient Educ Couns 102, 1558–1564 (2019). 10.1016/j.pec.2019.04.014

12 Lerman, C. et al. BRCA1 testing in families with hereditary breast-ovarian cancer. A prospective study of patient decision making and outcomes. JAMA 275, 1885–1892 (1996). doi:10.1001/jama.1996.03530480027036

13 Kelly, K. et al. Cancer genetics knowledge and beliefs and receipt of results in Ashkenazi Jewish individuals receiving counseling for BRCA1/2 mutations. Cancer Control 11, 236–244 (2004). 10.1177/107327480401100405

14 Kaphingst, K. A. et al. Patients’ understanding of and responses to multiplex genetic susceptibility test results. Genet Med 14, 681–687 (2012). 10.1038/gim.2012.22

15 Bradbury, A. R. et al. Longitudinal outcomes with cancer multigene panel testing in previously tested BRCA1/2 negative patients. Clin Genet 97, 601–609 (2020). 10.1111/cge.13716

16 Solomons, N. M., Lamb, A. E., Lucas, F. L., McDonald, E. F. & Miesfeldt, S. Examination of the Patient-Focused Impact of Cancer Telegenetics Among a Rural Population: Comparison with Traditional In-Person Services. Telemed J E Health 24, 130–138 (2018). 10.1089/tmj.2017.0073

17 Yanes, T. et al. Breast cancer polygenic risk scores: a 12-month prospective study of patient reported outcomes and risk management behavior. Genet Med 23, 2316–2323 (2021). 10.1038/s41436-021-01288-6

18 Keller, M. et al. Psychosocial outcome following genetic risk counselling for familial colorectal cancer. A comparison of affected patients and family members. Clin Genet 74, 414–424 (2008). 10.1111/j.1399-0004.2008.01089.x

19 Lemke, A. A. et al. Patient-Reported Outcomes and Experiences with Population Genetic Testing Offered Through a Primary Care Network. Genet Test Mol Biomarkers 25, 152–160 (2021). 10.1089/gtmb.2020.0275

20 Loeb, S. et al. Technology-enhanced AcceleRation of Germline Evaluation for Therapy (TARGET): A randomized controlled trial of a pretest patient-driven webtool vs. genetic counseling for prostate cancer germline testing. Contemp Clin Trials 119, 106821 (2022). 10.1016/j.cct.2022.106821

21 Underhill-Blazey, M. et al. Informing models of cancer genetics care in the era of multigene panel testing with patient-led recommendations. J Genet Couns 30, 268–282 (2021). 10.1002/jgc4.1317

22 Armstrong, J. et al. American BRCA Outcomes and Utilization of Testing (ABOUT) study: a pragmatic research model that incorporates personalized medicine/patient-centered outcomes in a real world setting. J Genet Couns 24, 18–28 (2015). 10.1007/s10897-014-9750-3

23 Hall, M. J., et al. Use and Patient-Reported Outcomes of Clinical Multigene Panel Testing for Cancer Susceptibility in the Multicenter Communication of Genetic Test Results by Telephone Study. JCO Precis Oncol 2 (2018). 10.1200/PO.18.00199

24 Hooker, G. W. et al. Longitudinal changes in patient distress following interactive decision aid use among BRCA1/2 carriers: a randomized trial. Med Decis Making 31, 412–421 (2011). 10.1177/0272989X10381283

25 Interrante, M. K. et al. Randomized Noninferiority Trial of Telephone vs In-Person Genetic Counseling for Hereditary Breast and Ovarian Cancer: A 12-Month Follow-Up. JNCI Cancer Spectr 1, pkx002 (2017). 10.1093/jncics/pkx002

26 Oberguggenberger, A. et al. Psychosocial outcomes and counselee satisfaction following genetic counseling for hereditary breast and ovarian cancer: A patient-reported outcome study. J Psychosom Res 89, 39–45 (2016). 10.1016/j.jpsychores.2016.08.005

27 Sie, A. S., Prins, J. B., Spruijt, L., Kets, C. M. & Hoogerbrugge, N. Can we test for hereditary cancer at 18 years when we start surveillance at 25? Patient reported outcomes. Fam Cancer 12, 675–682 (2013). 10.1007/s10689-013-9644-9

28 Makhnoon, S. et al. The FamilyTalk randomized controlled trial: patient-reported outcomes in clinical genetic sequencing for colorectal cancer. Cancer Causes Control 32, 483–492 (2021). 10.1007/s10552-021-01398-1

29 Eijzenga, W. et al. Routine assessment of psychosocial problems after cancer genetic counseling: results from a randomized controlled trial. Clin Genet 87, 419–427 (2015). 10.1111/cge.12473

30 Wing, S. E. et al. Recall of Genomic Testing Results Among Patients with Cancer. Oncologist 26, e2302–e2305 (2021). 10.1002/onco.13928

31 Kukafka, R. et al. Patient and Clinician Decision Support to Increase Genetic Counseling for Hereditary Breast and Ovarian Cancer Syndrome in Primary Care: A Cluster Randomized Clinical Trial. JAMA Netw Open 5, e2222092 (2022). 10.1001/jamanetworkopen.2022.22092

32 Peshkin, B. N. et al. Patient Perceptions of Telephone vs. In-Person BRCA1/BRCA2 Genetic Counseling. J Genet Couns 25, 472–482 (2016). 10.1007/s10897-015-9897-6

33 Russo, J. et al. Pretest Genetic Education Video Versus Genetic Counseling for Men Considering Prostate Cancer Germline Testing: A Patient-Choice Study to Address Urgent Practice Needs. JCO Precis Oncol 5 (2021). 10.1200/PO.21.00238

34 Audrain, J. et al. Psychological distress in women seeking genetic counseling for breast-ovarian cancer risk: the contributions of personality and appraisal. Ann Behav Med 19, 370–377 (1997). 10.1007/BF02895156

35 Hamilton, J. G. et al. Uptake and acceptability of a mainstreaming model of hereditary cancer multigene panel testing among patients with ovarian, pancreatic, and prostate cancer. Genet Med 23, 2105–2113 (2021). 10.1038/s41436-021-01262-2

36 Zigmond, A. S. & Snaith, R. P. The hospital anxiety and depression scale. Acta Psychiatr Scand 67, 361–370 (1983). 10.1111/j.1600-0447.1983.tb09716.x

37 Lerman, C. et al. Psychological side effects of breast cancer screening. Health Psychol 10, 259–267 (1991). 10.1037//0278-6133.10.4.259

38 Eijzenga, W. et al. Effect of routine assessment of specific psychosocial problems on personalized communication, counselors’ awareness, and distress levels in cancer genetic counseling practice: a randomized controlled trial. J Clin Oncol 32, 2998–3004 (2014). 10.1200/JCO.2014.55.4576

39 Horowitz, M., N. Wilner, and W. Alvarez. Impact of Event Scale: a measure of subjective stress. Psychosom Med 41, 209–218 (1979). 10.1097/00006842-197905000-00004

40 State Insurance Regulatory Authority: Guidelines for the management of acute whiplash-associated disorders – for health professionals. 3rd Edition edn, 47 (2014).

41 O’Connor, A. M. Validation of a decisional conflict scale. Med Decis Making 15, 25–30 (1995). 10.1177/0272989X9501500105

42 DeMarco, T. A., Peshkin, B. N., Mars, B. D. & Tercyak, K. P. Patient satisfaction with cancer genetic counseling: a psychometric analysis of the Genetic Counseling Satisfaction Scale. J Genet Couns 13, 293–304 (2004). 10.1023/b:jogc.0000035523.96133.bc

43 Hall, R., Bancroft, E., Pashayan, N., Kote-Jarai, Z. & Eeles, R. A. Genetics of prostate cancer: a review of latest evidence. J Med Genet 61, 915–926 (2024). 10.1136/jmg-2024-109845

44 Lai-Kwon, J., Yin, Z., Minchom, A. & Yap, C. Trends in patient-reported outcome use in early phase dose-finding oncology trials - an analysis of ClinicalTrials.gov. Cancer Med 10, 7943–7957 (2021). 10.1002/cam4.4307

45 The Genetic Information Non-Discrimination Act, Public Law 110–223, <http://www.gpo.gov/fdsys/pkg/PLAW-110publ233/pdf/PLAW-110publ233.pdf> (

46 Charter of Fundamental Rights of the European Union, <https://www.europarl.europa.eu/charter/pdf/text_en.pdf> (

47 Patrinos, D., Ghaly, M., Al-Shafai, M. & Zawati, M. H. Legal approaches to risk of harm in genetic counseling: perspectives from Quebec and Qatar. Front Genet 14, 1190421 (2023). 10.3389/fgene.2023.1190421

48 Luke, J. et al. Investigating disparity in access to Australian clinical genetic health services for Aboriginal and Torres Strait Islander people. Nat Commun 13, 4966 (2022). 10.1038/s41467-022-32707-0

49 Ascencio-Carbajal, T., Saruwatari-Zavala, G., Navarro-Garcia, F. & Frixione, E. Genetic/genomic testing: defining the parameters for ethical, legal and social implications (ELSI). BMC Med Ethics 22, 156 (2021). 10.1186/s12910-021-00720-5

50 Liddicoat, J. et al. Continental drift? Do European clinical genetic testing laboratories have a patent problem? Eur J Hum Genet 27, 997–1007 (2019). 10.1038/s41431-019-0368-7

51 Subhan, M. A. et al. Recent Advances with Precision Medicine Treatment for Breast Cancer including Triple-Negative Sub-Type. Cancers (Basel) 15 (2023). 10.3390/cancers15082204

52 Mucci, L. A. et al. Familial Risk and Heritability of Cancer Among Twins in Nordic Countries. JAMA 315, 68–76 (2016). 10.1001/jama.2015.17703

53 Moller, S. et al. The Heritability of Breast Cancer among Women in the Nordic Twin Study of Cancer. Cancer Epidemiol Biomarkers Prev 25, 145–150 (2016). 10.1158/1055-9965.EPI-15-0913

54 Harper, J. B., Greenberg, S. E., Hunt, T. C., Cooney, K. A. & O’Neil, B. B. Initial outcomes and insights from a novel high-risk prostate cancer screening clinic. Prostate 83, 151–157 (2023). 10.1002/pros.24447

55 Hendricks-Sturrup, R. M., Joseph, L. & Lu, C. Y. Patient-Reported Outcomes following Genetic Testing for Familial Hypercholesterolemia, Breast and Ovarian Cancer Syndrome, and Lynch Syndrome: A Systematic Review. J Pers Med 11 (2021). 10.3390/jpm11090850

56 Kalra, A. et al. Uptake and patient-related outcomes of mainstreaming genetic testing: a systematic review and meta-analysis. Am J Obstet Gynecol (2025). 10.1016/j.ajog.2025.04.044

57 Ringwald, J. et al. Psychological Distress, Anxiety, and Depression of Cancer-Affected BRCA1/2 Mutation Carriers: a Systematic Review. J Genet Couns 25, 880–891 (2016). 10.1007/s10897-016-9949-6

58 Heshka, J. T., Palleschi, C., Howley, H., Wilson, B. & Wells, P. S. A systematic review of perceived risks, psychological and behavioral impacts of genetic testing. Genet Med 10, 19–32 (2008). 10.1097/GIM.0b013e31815f524f

59 Alwaal, A., Awad, M., Boggs, N., Kuzbel, J. & Snoad, B. Sexual Health Inventory for Men Questionnaire as a Screening Method for Erectile Dysfunction in a General Urology Clinic. Sex Med 8, 660–663 (2020). 10.1016/j.esxm.2020.08.002

60 Vriesen, N., Carmany, E. P. & Natoli, J. L. Clinical outcomes of preimplantation genetic testing for hereditary cancer syndromes: A systematic review. Prenat Diagn 42, 201–211 (2022). 10.1002/pd.6084

61 Yanes, T., Willis, A. M., Meiser, B., Tucker, K. M. & Best, M. Psychosocial and behavioral outcomes of genomic testing in cancer: a systematic review. Eur J Hum Genet 27, 28–35 (2019). 10.1038/s41431-018-0257-5

62 Minvielle, E. et al. The use of patient-reported outcome and experience measures for health policy purposes: A scoping review in oncology. Health Policy 129, 104702 (2023). 10.1016/j.healthpol.2022.12.010

63 Mokkink, L. B. et al. The COSMIN study reached international consensus on taxonomy, terminology, and definitions of measurement properties for health-related patient-reported outcomes. J Clin Epidemiol 63, 737–745 (2010). 10.1016/j.jclinepi.2010.02.006

64 Terwee, C. B. et al. COSMIN methodology for evaluating the content validity of patient-reported outcome measures: a Delphi study. Qual Life Res 27, 1159–1170 (2018). 10.1007/s11136-018-1829-0

65 Moher, D. et al. What contributions do languages other than English make on the results of meta-analyses? J Clin Epidemiol 53, 964–972 (2000). 10.1016/s0895-4356(00)00188-8

66 Rothwell, E. et al. Patient outcomes associated with group and individual genetic counseling formats. Fam Cancer 11, 97–106 (2012). 10.1007/s10689-011-9486-2

67 McCuaig, J. M. et al. Reflex BRCA1 and BRCA2 tumour genetic testing for high-grade serous ovarian cancer: streamlined for clinicians but what do patients think? Hered Cancer Clin Pract 20, 15 (2022). 10.1186/s13053-022-00221-5

68 Patrick-Miller, L. et al. Implementation and outcomes of telephone disclosure of clinical BRCA1/2 test results. Patient Educ Couns 93, 413–419 (2013). 10.1016/j.pec.2013.08.009

69 Bradbury, A. R. et al. Randomized Noninferiority Trial of Telephone vs In-Person Disclosure of Germline Cancer Genetic Test Results. J Natl Cancer Inst 110, 985–993 (2018). 10.1093/jnci/djy015

70 Blanchette, P. S. et al. Genomic testing in cancer: patient knowledge, attitudes, and expectations. Cancer 120, 3066–3073 (2014). 10.1002/cncr.28807

71 Langer, M. M. et al. Development and Validation of a Genomic Knowledge Scale to Advance Informed Decision Making Research in Genomic Sequencing. MDM Policy Pract 2 (2017). 10.1177/2381468317692582

72 Hamilton, J. G. et al. Maternal perceptions of BRCA genetic counseling communication processes about disclosing cancer risk information to children and adult relatives. Psychooncology 27, 1825–1832 (2018). 10.1002/pon.4733

73 De Silva, D. L. et al. Universal genetic testing for women with newly diagnosed breast cancer in the context of multidisciplinary team care. Med J Aust 218, 368–373 (2023). 10.5694/mja2.51906

74 Armstrong, J. et al. Utilization and Outcomes of BRCA Genetic Testing and Counseling in a National Commercially Insured Population: The ABOUT Study. JAMA Oncol 1, 1251–1260 (2015). 10.1001/jamaoncol.2015.3048

75 Berkenstadt, M., Shiloh, S., Barkai, G., Katznelson, M. B. & Goldman, B. Perceived personal control (PPC): a new concept in measuring outcome of genetic counseling. Am J Med Genet 82, 53–59 (1999). 10.1002/(sici)1096-8628(19990101)82:1

76 Harris, J. N. et al. Interest in genetic testing among affected men from hereditary prostate cancer families and their unaffected male relatives. Genet Med 11, 344–355 (2009). 10.1097/GIM.0b013e31819b2425

77 McCuaig, J. M. et al. A Comparison of Patient-Reported Outcomes Following Consent for Genetic Testing Using an Oncologist- or Genetic Counselor-Mediated Model of Care. Curr Oncol 28, 1459–1471 (2021). 10.3390/curroncol28020138

78 McCuaig, J. M. et al. Patient-reported outcomes associated with reflex BRCA1/2 tumor and subsequent germline panel genetic testing for high-grade serous ovarian cancer. J Genet Couns 32, 503–513 (2023). 10.1002/jgc4.1661

79 Kasparian, N. A., Meiser, B., Butow, P. N., Simpson, J. M. & Mann, G. J. Genetic testing for melanoma risk: a prospective cohort study of uptake and outcomes among Australian families. Genet Med 11, 265–278 (2009). 10.1097/GIM.0b013e3181993175

80 George, A. et al. Implementing rapid, robust, cost-effective, patient-centred, routine genetic testing in ovarian cancer patients. Sci Rep 6, 29506 (2016). 10.1038/srep29506

81 Kemp, Z. et al. Evaluation of Cancer-Based Criteria for Use in Mainstream BRCA1 and BRCA2 Genetic Testing in Patients With Breast Cancer. JAMA Netw Open 2, e194428 (2019). 10.1001/jamanetworkopen.2019.4428

82 Richardson, M. et al. Oncology Clinic-Based Hereditary Cancer Genetic Testing in a Population-Based Health Care System. Cancers (Basel) 12 (2020). 10.3390/cancers12020338

83 Cella, D. et al. A brief assessment of concerns associated with genetic testing for cancer: the Multidimensional Impact of Cancer Risk Assessment (MICRA) questionnaire. Health Psychol 21, 564–572 (2002). 10.1037/0278-6133.21.6.564

84 Pieterse, A. H., van Dulmen, A. M., Beemer, F. A., Bensing, J. M. & Ausems, M. G. Cancer genetic counseling: communication and counselees’ post-visit satisfaction, cognitions, anxiety, and needs fulfillment. J Genet Couns 16, 85–96 (2007). 10.1007/s10897-006-9048-1

85 Lemke, A. A. et al. Patient perspectives following pharmacogenomics results disclosure in an integrated health system. Pharmacogenomics 19, 321–331 (2018). 10.2217/pgs-2017-0191

86 Li, M. et al. The Feelings About genomiC Testing Results (FACToR) Questionnaire: Development and Preliminary Validation. J Genet Couns 28, 477–490 (2019). 10.1007/s10897-018-0286-9

87 Zoltick, E. S. et al. Predispositional genome sequencing in healthy adults: design, participant characteristics, and early outcomes of the PeopleSeq Consortium. Genome Med 11, 10 (2019). 10.1186/s13073-019-0619-9

88 Finlay, E. et al. Factors determining dissemination of results and uptake of genetic testing in families with known BRCA1/2 mutations. Genet Test 12, 81–91 (2008). 10.1089/gte.2007.0037

89 Patenaude, A. F. et al. Sharing BRCA1/2 test results with first-degree relatives: factors predicting who women tell. J Clin Oncol 24, 700–706 (2006). 10.1200/JCO.2005.01.7541

90 Schwartz, L. M., Woloshin, S., Black, W. C. & Welch, H. G. The role of numeracy in understanding the benefit of screening mammography. Ann Intern Med 127, 966–972 (1997). 10.7326/0003-4819-127-11-199712010-00003

91 Fagerlin, A. et al. Measuring numeracy without a math test: development of the Subjective Numeracy Scale. Med Decis Making 27, 672–680 (2007). 10.1177/0272989X07304449

92 Zikmund-Fisher, B. J., Smith, D. M., Ubel, P. A. & Fagerlin, A. Validation of the Subjective Numeracy Scale: effects of low numeracy on comprehension of risk communications and utility elicitations. Med Decis Making 27, 663–671 (2007). 10.1177/0272989X07303824

93 Norman, C. D. & Skinner, H. A. eHEALS: The eHealth Literacy Scale. J Med Internet Res 8, e27 (2006). 10.2196/jmir.8.4.e27

94 Chew, L. D., Bradley, K. A. & Boyko, E. J. Brief questions to identify patients with inadequate health literacy. Fam Med 36, 588–594 (2004).

95 Rockhill, B., Spiegelman, D., Byrne, C., Hunter, D. J. & Colditz, G. A. Validation of the Gail et al. model of breast cancer risk prediction and implications for chemoprevention. J Natl Cancer Inst 93, 358–366 (2001). 10.1093/jnci/93.5.358

96 Gail, M. H. et al. Projecting individualized probabilities of developing breast cancer for white females who are being examined annually. J Natl Cancer Inst 81, 1879–1886 (1989). 10.1093/jnci/81.24.1879

97 Tyrer, J., Duffy, S. W. & Cuzick, J. A breast cancer prediction model incorporating familial and personal risk factors. Stat Med 23, 1111–1130 (2004). 10.1002/sim.1668

98 Schwartz, M. D. et al. Bilateral prophylactic oophorectomy and ovarian cancer screening following BRCA1/BRCA2 mutation testing. J Clin Oncol 21, 4034–4041 (2003). 10.1200/JCO.2003.01.088

99 Morman, N. A., Byrne, L., Collins, C., Reynolds, K. & Bell, J. G. Breast Cancer Risk Assessment at the Time of Screening Mammography: Perceptions and Clinical Management Outcomes for Women at High Risk. J Genet Couns 26, 776–784 (2017). 10.1007/s10897-016-0050-y

100 Codori, A. M. et al. Attitudes toward colon cancer gene testing: factors predicting test uptake. Cancer Epidemiol Biomarkers Prev 8, 345–351 (1999).

101 Evans, C. N. et al. Impact of genomic testing and patient-reported outcomes on receipt of adjuvant chemotherapy. Breast Cancer Res Treat 156, 549–555 (2016). 10.1007/s10549-016-3780-5

102 Ware, J. E., Jr. & Sherbourne, C. D. The MOS 36-item short-form health survey (SF-36). I. Conceptual framework and item selection. Med Care 30, 473–483 (1992).

103 Ware, J., Jr., Kosinski, M. & Keller, S. D. A 12-Item Short-Form Health Survey: construction of scales and preliminary tests of reliability and validity. Med Care 34, 220–233 (1996). 10.1097/00005650-199603000-00003

104 Aaronson, N. K. et al. The European Organization for Research and Treatment of Cancer QLQ-C30: a quality-of-life instrument for use in international clinical trials in oncology. J Natl Cancer Inst 85, 365–376 (1993). 10.1093/jnci/85.5.365

105 Oken, M. M. et al. Toxicity and response criteria of the Eastern Cooperative Oncology Group. Am J Clin Oncol 5, 649–655 (1982).

106 Rosen, R. C., Cappelleri, J. C., Smith, M. D., Lipsky, J. & Pena, B. M. Development and evaluation of an abridged, 5-item version of the International Index of Erectile Function (IIEF-5) as a diagnostic tool for erectile dysfunction. Int J Impot Res 11, 319–326 (1999). 10.1038/sj.ijir.3900472

107 Barry, M. J. et al. The American Urological Association symptom index for benign prostatic hyperplasia. The Measurement Committee of the American Urological Association. J Urol 148, 1549–1557; discussion 1564 (1992). 10.1016/s0022-5347(17)36966-5

108 Gomez, R., Stavropoulos, V., Zarate, D. & Palikara, O. Symptom Checklist-90-Revised: A structural examination in relation to family functioning. PLoS One 16, e0247902 (2021). 10.1371/journal.pone.0247902

109 Derogatis, L. R., Lipman, R. S. & Covi, L. SCL-90: an outpatient psychiatric rating scale--preliminary report. Psychopharmacol Bull 9, 13–28 (1973).

110 Derogatis, L. R. & Melisaratos, N. The Brief Symptom Inventory: an introductory report. Psychol Med 13, 595–605 (1983).

111 Adawi, M. et al. Psychometric properties of the Brief Symptom Inventory in nomophobic subjects: insights from preliminary confirmatory factor, exploratory factor, and clustering analyses in a sample of healthy Italian volunteers. Psychol Res Behav Manag 12, 145–154 (2019). 10.2147/PRBM.S173282

112 Braehler W, S. J. The Giessen Complaints Inventory (German). (1995).

113 Lovibond, P. F. & Lovibond, S. H. The structure of negative emotional states: comparison of the Depression Anxiety Stress Scales (DASS) with the Beck Depression and Anxiety Inventories. Behav Res Ther 33, 335–343 (1995). 10.1016/0005-7967(94)00075-u

114 Lovibond, S. H. L., P.F. Manual for the Depression Anxiety Stress Scales. 2nd Ed edn, (1995).

115 Henry, J. D. & Crawford, J. R. The short-form version of the Depression Anxiety Stress Scales (DASS-21): construct validity and normative data in a large non-clinical sample. Br J Clin Psychol 44, 227–239 (2005). 10.1348/014466505X29657

116 Touzani, R. et al. Adaptation and validation of the memorial anxiety scale for prostate cancer (MAX-PC) in a sample of French men. J Patient Rep Outcomes 3, 60 (2019). 10.1186/s41687-019-0150-1

117 Roth, A. J. et al. The memorial anxiety scale for prostate cancer: validation of a new scale to measure anxiety in men with with prostate cancer. Cancer 97, 2910–2918 (2003). 10.1002/cncr.11386

118 Zsido, A. N., Teleki, S. A., Csokasi, K., Rozsa, S. & Bandi, S. A. Development of the short version of the spielberger state-trait anxiety inventory. Psychiatry Res 291, 113223 (2020). 10.1016/j.psychres.2020.113223

119 GA, S. C. G. R. L. R. V. P. J. Manual for the State-Trait Anxiety Inventory (Form Y). (Mind Garden, Inc, 1983).

120 Spitzer, R. L., Kroenke, K., Williams, J. B. & Lowe, B. A brief measure for assessing generalized anxiety disorder: the GAD-7. Arch Intern Med 166, 1092–1097 (2006). 10.1001/archinte.166.10.1092

121 Kroenke, K., Spitzer, R. L., Williams, J. B. & Lowe, B. An ultra-brief screening scale for anxiety and depression: the PHQ-4. Psychosomatics 50, 613–621 (2009). 10.1176/appi.psy.50.6.613

122 Cohen, S., Kamarck, T. & Mermelstein, R. A global measure of perceived stress. J Health Soc Behav 24, 385–396 (1983).

123 Cohen, S. a. W., G. Perceived Stress in a Probability Sample of the United States. ( The Social Psychology of Health, 1988).

124 Dibble, K. E., Donorfio, L. K. M., Britner, P. A. & Bellizzi, K. M. Stress, anxiety, and health-related quality of life in BRCA1/2-positive women with and without cancer: A comparison of four US female samples. Gynecol Oncol Rep 42, 101033 (2022). 10.1016/j.gore.2022.101033

125 Thewes, B., Zachariae, R., Christensen, S., Nielsen, T. & Butow, P. The Concerns About Recurrence Questionnaire: validation of a brief measure of fear of cancer recurrence amongst Danish and Australian breast cancer survivors. J Cancer Surviv 9, 68–79 (2015). 10.1007/s11764-014-0383-1

126 Katz, S. J. et al. Association of Germline Genetic Test Type and Results With Patient Cancer Worry After Diagnosis of Breast Cancer. JCO Precis Oncol 2018 (2018). 10.1200/PO.18.00225

127 Schwartz, M. D., Taylor, K. L. & Willard, K. S. Prospective association between distress and mammography utilization among women with a family history of breast cancer. J Behav Med 26, 105–117 (2003). 10.1023/a:1023078521319

128 Ownby, K. K. Use of the Distress Thermometer in Clinical Practice. J Adv Pract Oncol 10, 175–179 (2019).

129 National Comprehensive Cancer, N. Distress management. Clinical practice guidelines. J Natl Compr Canc Netw 1, 344–374 (2003). 10.6004/jnccn.2003.0031

130 Weiss, D. S., & Marmar, C. R.. The Impact of Event Scale - Revised. 399–411 (Assessing psychological trauma and PTSD, 1996).

131 Kautz-Freimuth, S. et al. Evaluation of two evidence-based decision aids for female BRCA1/2 mutation carriers in Germany: study protocol for a randomised controlled parallel-group trial. Trials 23, 157 (2022). 10.1186/s13063-022-06081-7

132 Holmes-Rovner, M. et al. Patient satisfaction with health care decisions: the satisfaction with decision scale. Med Decis Making 16, 58–64 (1996). 10.1177/0272989X9601600114

133 O’Connor, A. (1995).

134 Cranney, A. et al. Development and pilot testing of a decision aid for postmenopausal women with osteoporosis. Patient Educ Couns 47, 245–255 (2002). 10.1016/s0738-3991(01)00218-x

135 Brehaut, J. C. et al. Validation of a decision regret scale. Med Decis Making 23, 281–292 (2003). 10.1177/0272989X03256005

136 Degner, L. F., Sloan, J. A. & Venkatesh, P. The Control Preferences Scale. Can J Nurs Res 29, 21–43 (1997).

137 McAllister, M., Wood, A. M., Dunn, G., Shiloh, S. & Todd, C. The Genetic Counseling Outcome Scale: a new patient-reported outcome measure for clinical genetics services. Clin Genet 79, 413–424 (2011). 10.1111/j.1399-0004.2011.01636.x

138 Pieterse, A. et al. QUOTE-gene(ca): development of a counselee-centered instrument to measure needs and preferences in genetic counseling for hereditary cancer. Psychooncology 14, 361–375 (2005). 10.1002/pon.853

139 Salemink, S. et al. Focusing on patient needs and preferences may improve genetic counseling for colorectal cancer. J Genet Couns 22, 118–124 (2013). 10.1007/s10897-012-9519-5

140 Shiloh, S., Avdor, O. & Goodman, R. M. Satisfaction with genetic counseling: dimensions and measurement. Am J Med Genet 37, 522–529 (1990). 10.1002/ajmg.1320370419

141 Bjorvatn, C., Eide, G. E., Hanestad, B. R., Hamang, A. & Havik, O. E. Intrusion and avoidance in subjects undergoing genetic investigation and counseling for hereditary cancer. Support Care Cancer 17, 1371–1381 (2009). 10.1007/s00520-009-0594-6

142 Nordin, K., Liden, A., Hansson, M., Rosenquist, R. & Berglund, G. Coping style, psychological distress, risk perception, and satisfaction in subjects attending genetic counselling for hereditary cancer. J Med Genet 39, 689–694 (2002). 10.1136/jmg.39.9.689

