## Supplementary Material for "Genetic testing in cancer: a systematic review of patient reported measures"

### Supplementary Table 1. Overview of the other PRMs identified by the systematic literature search.

| Domains/PRMs/Tools (Abbreviation) | Information about tool | Original Development Paper/Adaptation | Number of Studies |
|---|---|---|---|
| *Perceptions* | | | |
| BREAST-Q | Focus on perception of breast surgery outcomes. Scales for augmentation, reduction, reconstruction, and process of care[1]. | Pusic *et al.,* 2006[1]. Appears to be used without adaptation in this study. | n=1 (van Egdom *et al.,* 2020[2]) |
| Perceptions of chemotherapy | Assessed the impact of genomic testing on receipt of adjuvant chemotherapy for their breast cancer. Patients were asked to rate pros (n=8) and cons (n=8) of receiving chemotherapy. | Adapted from previously used measures by O'Neil *et al.,* 2007[3] and O'Neil *et al.,* 2010[4]. | n=1 (Evans *et al.,* 2016[5]) |
| *Telemedicine* | | | |
| Perception and experience | Open-ended questions; appears study-specific. | Adapted from Patrick-Miller *et al.,* 2013[6] & 2014[7]. | n=1 (Bradbury *et al.,* 2016[8]) |
| Telemedicine Satisfaction Questionnaire | Developed for paediatric care, aimed at families: covers comfort, comparison to face to face visit, privacy concerns, technical difficulties, satisfaction. | 13-items adapted from Dick *et al.,* 1999[9]. Evaluated patient-perceived provider comfort, patient satisfaction with privacy, and patient comfort with audio/visual technology. | n=1 (Bradbury *et al.*, 2016[8]) |
| Telemedicine Satisfaction Questionnaire | Satisfaction with telemedicine; covers quality of care, perception of encounter, similarity of telemedicine encounter with in-person encounter[10]. 6-items on a 4-point Likert scale. | Five of the six items were adapted from the validated Telemedicine Satisfaction Questionnaire[10]. The confidentiality question was adapted from a similar item in the Utah Telehealth Network's Patient Satisfaction Survey (https://healthit.ahrq.gov/health-it-tools-and-resources/health-it-survey-compendium/utah-telehealth-network-telehealth-patient). | n=1 (Solomons *et al.,* 2018[11]) |
| Satisfaction and acceptability of telephone counselling session | Study-specific measure that assessed the counselees' satisfaction and perceived acceptability, in terms of usefulness, of the extra telephone session. | Appears to be used without validation in this study. | n=1 (Eijzenga *et al.,* 2015[12]) |
| *Other* | | | |
| Threatening Medical Situations Inventory | Cognitive response to medical threat[13]. | A shortened version using two situations was used here. This version has been previously validated to assess coping tyles in hypothetical medical situations[13-15]. | n=1 (Sie *et al.,* 2013[16]) |

| | | | |
|---|---|---|---|
| Interpersonal Support Evaluation List (ISEL) | Perceived social support[17]. 30 items on a four-point scale, with the average sum score used, as previously described[18]. | King *et al.,* 1993[18]. Appears to be used without adaptation in this study. | n=1 (Bjorvatn *et al.,* 2009[19]) |
| Ease of access to services | Five questions focused on economic burden, distance travelled, distance travelled, etc. | Adapted from a Maine Medical Center Survey used to assess ongoing general (noncancer) telegenetic care[11]. | n=1 (Solomons *et al.,* 2018[11]) |
| Revised Life Orientation Test (LOT-R) | Evaluation of dispositional optimism; 10-item scale, assessing optimism and pessimism. | Scheier *et al.,* 1994[20]. Appears to be used without adaptation in this study. Internal consistency of the scale was determined in their cohort (α=0.79). | n=1 (McCuaig *et al.,* 2022[21]) |
| Parent-Adolescent Communication Scale (PACS) | Measures quality of the parent-child communication relationship (20 items). | Olson & Barnes *et al.,* 1982[22]. Appears to be used without adaptation in this study. They also used an 11-item Pediatric *BRCA* Testing Attitudes Scale (adapted from Peshkin *et al.,* 2009[23]) to understand pediatric genetic testing attitudes after mothers of minor-age children received *BRCA* results. | n=1 (Hamilton *et al.,* 2018[24]) |

### REFERENCES


1 Pusic, A. L. *et al.* Development of a new patient-reported outcome measure for breast surgery: the BREAST-Q. *Plast Reconstr Surg* **124**, 345-353 (2009). https://doi.org:10.1097/PRS.0b013e3181aee807

2 van Egdom, L. S. E. *et al.* Patient-Reported Outcome Measures may optimize shared decision-making for cancer risk management in BRCA mutation carriers. *Breast Cancer* **27**, 426-434 (2020). https://doi.org:10.1007/s12282-019-01033-7

3 O'Neill, S. C. *et al.* Women's interest in gene expression analysis for breast cancer recurrence risk. *J Clin Oncol* **25**, 4628-4634 (2007). https://doi.org:10.1200/JCO.2006.09.6255

4 O'Neill, S. C. *et al.* BRCA1/2 test results impact risk management attitudes, intentions, and uptake. *Breast Cancer Res Treat* **124**, 755-764 (2010). https://doi.org:10.1007/s10549-010-0881-4

5 Evans, C. N. *et al.* Impact of genomic testing and patient-reported outcomes on receipt of adjuvant chemotherapy. *Breast Cancer Res Treat* **156**, 549-555 (2016). https://doi.org:10.1007/s10549-016-3780-5

6 Patrick-Miller, L. *et al.* Implementation and outcomes of telephone disclosure of clinical BRCA1/2 test results. *Patient Educ Couns* **93**, 413-419 (2013). https://doi.org:10.1016/j.pec.2013.08.009

7 Patrick-Miller, L. J. *et al.* Development of a communication protocol for telephone disclosure of genetic test results for cancer predisposition. *JMIR Res Protoc* **3**, e49 (2014). https://doi.org:10.2196/resprot.3337

8 Bradbury, A. *et al.* Utilizing Remote Real-Time Videoconferencing to Expand Access to Cancer Genetic Services in Community Practices: A Multicenter Feasibility Study. *J Med Internet Res* **18**, e23 (2016). https://doi.org:10.2196/jmir.4564

9 Dick, P. T., Filler, R. & Pavan, A. Participant satisfaction and comfort with multidisciplinary pediatric telemedicine consultations. *J Pediatr Surg* **34**, 137-141; discussion 141-132 (1999). https://doi.org:10.1016/s0022-3468(99)90244-0

10 Yip, M. P., Chang, A. M., Chan, J. & MacKenzie, A. E. Development of the Telemedicine Satisfaction Questionnaire to evaluate patient satisfaction with telemedicine: a preliminary study. *J Telemed Telecare* **9**, 46-50 (2003). https://doi.org:10.1258/135763303321159693

11 Solomons, N. M., Lamb, A. E., Lucas, F. L., McDonald, E. F. & Miesfeldt, S. Examination of the Patient-Focused Impact of Cancer Telegenetics Among a Rural Population: Comparison with Traditional In-Person Services. *Telemed J E Health* **24**, 130-138 (2018). https://doi.org:10.1089/tmj.2017.0073

12 Eijzenga, W. *et al.* Routine assessment of psychosocial problems after cancer genetic counseling: results from a randomized controlled trial. *Clin Genet* **87**, 419-427 (2015). https://doi.org:10.1111/cge.12473

13 van Zuuren FJ, d. G. K., Mulder NL, Muris P. Coping with medical threat: An evaluation of the Threatening Medical Situations Inventory (TMSI). *Personality and Individual Differences* **21**, 21-31 (1996). https://doi.org:https://doi.org/10.1016/0191-8869(96)00029-3

14 Ong, L. M. *et al.* Cancer patients' coping styles and doctor-patient communication. *Psychooncology* **8**, 155-166 (1999). https://doi.org:10.1002/(SICI)1099-1611(199903/04)8:2<155::AID-PON350>3.0.CO;2-A

15 Timmermans, L. M., van Zuuren, F. J., van der Maazen, R. W., Leer, J. W. & Kraaimaat, F. W. Monitoring and blunting in palliative and curative radiotherapy consultations. *Psychooncology* **16**, 1111-1120 (2007). https://doi.org:10.1002/pon.1177

16 Sie, A. S., Prins, J. B., Spruijt, L., Kets, C. M. & Hoogerbrugge, N. Can we test for hereditary cancer at 18 years when we start surveillance at 25? Patient reported outcomes. *Fam Cancer* **12**, 675-682 (2013). https://doi.org:10.1007/s10689-013-9644-9
17 Merz, E. L. *et al.* Validation of interpersonal support evaluation list-12 (ISEL-12) scores among English- and Spanish-speaking Hispanics/Latinos from the HCHS/SOL Sociocultural Ancillary Study. *Psychol Assess* **26**, 384-394 (2014). https://doi.org:10.1037/a0035248
18 King, K. B., Reis, H. T., Porter, L. A. & Norsen, L. H. Social support and long-term recovery from coronary artery surgery: effects on patients and spouses. *Health Psychol* **12**, 56-63 (1993). https://doi.org:10.1037//0278-6133.12.1.56
19 Bjorvatn, C., Eide, G. E., Hanestad, B. R., Hamang, A. & Havik, O. E. Intrusion and avoidance in subjects undergoing genetic investigation and counseling for hereditary cancer. *Support Care Cancer* **17**, 1371-1381 (2009). https://doi.org:10.1007/s00520-009-0594-6
20 Scheier, M. F., Carver, C. S. & Bridges, M. W. Distinguishing optimism from neuroticism (and trait anxiety, self-mastery, and self-esteem): a reevaluation of the Life Orientation Test. *J Pers Soc Psychol* **67**, 1063-1078 (1994). https://doi.org:10.1037//0022-3514.67.6.1063
21 McCuaig, J. M. *et al.* Reflex BRCA1 and BRCA2 tumour genetic testing for high-grade serous ovarian cancer: streamlined for clinicians but what do patients think? *Hered Cancer Clin Pract* **20**, 15 (2022). https://doi.org:10.1186/s13053-022-00221-5
22 Barnes HL, O. D. *Parent-Adolescent Communication Scale*. 33-48 (1982).
23 Peshkin, B. N. *et al.* Brief assessment of parents' attitudes toward testing minor children for hereditary breast/ovarian cancer genes: development and validation of the Pediatric BRCA1/2 Testing Attitudes Scale (P-TAS). *J Pediatr Psychol* **34**, 627-638 (2009). https://doi.org:10.1093/jpepsy/jsn033
24 Hamilton, J. G. *et al.* Maternal perceptions of BRCA genetic counseling communication processes about disclosing cancer risk information to children and adult relatives. *Psychooncology* **27**, 1825-1832 (2018). https://doi.org:10.1002/pon.4733
